# Two Separate, Large Cohorts Reveal Potential Modifiers of Age-Associated Variation in Visual Reaction Time Performance

**DOI:** 10.1101/2020.07.06.20147462

**Authors:** J.S. Talboom, M.D. De Both, M.A. Naymik, A.M. Schmidt, C.R. Lewis, W.M. Jepsen, A.K. Håberg, T. Rundek, B.E. Levin, S. Hoscheidt, Y. Bolla, R.D. Brinton, N.J. Schork, M. Hay, C.A. Barnes, E. Glisky, L. Ryan, M.J. Huentelman

## Abstract

To identify individual differences and potential factors influencing age-related cognitive decline and disease, we created MindCrowd. MindCrowd is a cross-sectional web-based assessment of simple visual (sv) reaction time (RT, index of processing speed) and paired-associate learning (PAL, index of verbal episodic memory). svRT and PAL results were combined with 22 survey questions. Analysis of MindCrowd’s svRT data revealed education and reported stroke as potential modifiers of changes in processing speed and memory from younger to older ages (*n*_*total*_ *=* 75,666, *n*_*women*_ = 47,700, *n*_men_ = 27,966; ages 18-85 years old, mean (*M*)_*Age*_ = 46.54, standard deviation *(SD)*_*Age*_ = 18.40). To complement this work, we evaluated complex recognition reaction time (cvrRT) in the UK Biobank cohort (*n*_*total*_ *=* 158,249 *n*_*women*_ = 89,333 *n*_men_ = 68,916; ages 40-70 years old, *M*_*Age*_ = 55.81, *SD*_*Age*_ = 7.72). Similarities between the UK Biobank and MindCrowd were assessed using a subset of the MindCrowd cohort. Labeled UKBb MindCrowd (*n*_*total*_ *=* 39,795, *n*_*women*_ = 29,640, *n*_men_ = 10,155; ages 40-70 years old, *M*_*Age*_ = 56.59, *SD*_*Age*_ = 8.16), this subset was carefully selected to mirror the UK Biobank. An identical linear model (LM) was used to assess both cohorts. The LM revealed similarities between MindCrowd and the UK Biobank across most results, despite obvious cohort differences (e.g., US vs. the UK). Notable divergent findings from the UK Biobank included (1) a first-degree family history of Alzheimer’s disease (FHAD) was associated with longer RTs in only. (2) Compared to being a man with more education, being a woman was associated with longer cvrRT length differences observed from younger to older ages. Divergent results from UKBb MindCrowd include more education and reported smoking. More education was associated with shorter and smoking longer cvrRTs differences observed from younger to older ages. Collected with our prior work, MindCrowd is beginning to reveal the intricate network connecting processing speed, memory, and cognition to healthy and pathological brain aging.

---

Reaction time (RT), an index of processing speed or efficiency in the central nervous system (CNS) ^1^, is an essential factor in higher cognitive function^2,3^ and is profoundly affected by age ^4^. In fact, of the studied demographics, age is the main factor known to influence RT^4^. Processing speed is an important limiting factor for most aspects of cognition during aging, most notably memory^5,6^. In studies where processing speed was used as a covariate, the age-related variance in various episodic memory measures was reduced or even eliminated^7,8^. Moreover, studies comparing varied factors and tests of age-related episodic memory deficit implicate age-related decline in processing speed as the main mediator^9-11^. These findings collectively suggest that RT is a useful index of age-related cognitive decline, healthy brain aging, and neurodevelopment.

RT can be operationally defined as “<u>simple,</u>” which typically involves a non-choice reaction to a <u>visual</u> stimulus (svRT). RT can also be operationally defined as “<u>complex</u>,” which involves a reaction to one or more <u>visual</u> stimuli after <u>recognition</u> (cvrRT) of correct stimuli and inhibiting incorrect stimuli ^12^. svRT demonstrates variability between individuals, which is akin to paired-associate learning (PAL) and is influenced by genetic and environmental factors^13^. In addition, svRT effects are well noted across the field of neurology; for example, AD and stroke patients show slower svRT and higher inter-individual variability ^14,15^. However, due to the limitations of traditional research methods, the body of work concerning RT examined only limited ranges of demographic, health, medical, and lifestyle factors in small cohorts. For example, prior work’s demographics consisted of college-aged students, well-educated older adults^16-18^, or athletes ^19-21^.

Further, with notable exceptions^22-24^, many studies had few participants (e.g., *n* <1000) and were therefore powered to detect only variables with large effect size and to lead to spurious non-replicable findings^25^. Consequently, many RT studies had minimal ability to reveal low frequency factors or those with subtle effect sizes and conduct more sophisticated analyses (e.g., ANOVA vs. Growth Modeling) to find interactions and moderators. Collectively, this suggests that if RT performance can inform models of disease or normative and atypical aging, we need a deeper understanding of the normal variation of RT and the genetic and environmental factors associated with RT performance.

This study aimed to characterize RT across a broad range of demographic, health, medical, and lifestyle variables commonly associated with cognitive performance and AD risk. To do this, we utilized both the MindCrowd and UK Biobank cohorts^26,27^, comprising over 233 thousand combined participants, to model RT as a function of 11 or more demographic, health, medical, and lifestyle factors. These factors have been previously associated with aging and cognition ^28-33^. Based on our prior work and earlier RT research^34^, we hypothesized that RT, via its structure of factor association and modifications, would reveal meaningful connections to healthy brain aging.

## Materials and Methods

### Study Participants

#### MindCrowd: Overview

In January 2013, we launched our internet-based study at www.mindcrowd.org. Website visitors 18 years or older were asked to consent to our study before any data collection via an electronic consent form. As of 3-26-2020, we have had 356,674 non-duplicate or distinct visitors to the website. Of these distinct visitors, over 194,542 (54%) consented to take part. The final data set had 75,666 (54% of consented individuals) participants who completed a simple visual reaction time (svRT) and paired-associate learning (PAL) tasks and answered 22 demographic, lifestyle, and health questions. Approval for this study was obtained from the Western Institutional Review Board (WIRB study number 1129241).

### MindCrowd: simple visual reaction time (svRT)

After consenting to the study and answering five demographic questions (i.e., age, biological sex, years of education, primary language, and country where they reside), participants were asked to complete a web-based svRT task. We chose svRT because it is a simple central and peripheral nervous system-dependent task influenced by intelligence and brain injury^35^. Participants were presented with a pink sphere that appeared at random intervals (between 1-10 seconds) on the screen, and they were instructed to respond as quickly as possible after the sphere appeared by pressing the enter/return key on their keyboard. Once the participant responded, the sphere disappeared until the next trial. Each participant received a total of five trials. The sphere stayed on the screen until the participant responded. The dependent variable, response time in milliseconds (msec), was recorded from the sphere’s appearance on the screen to the participant’s key press or screen touch.

### MindCrowd: paired-associate learning (PAL)

Next, participants were presented with the PAL task. For this cognitive task, during the learning phase, participants were shown 12 word-pairs, one word-pair at a time (2s/word-pair). During the recall phase, participants were given the first word of each pair and were asked to use their keyboard to type in (i.e., recall) the missing word. This learning-recall procedure was repeated for two more trials. Before beginning the task, each participant received one practice trial consisting of three word-pairs not contained in the 12 used during the test. Word-pairs were presented in different random orders during each learning and each recall phase. The same word pairs and order of presentation were used for all participants. The dependent variable/criterion was the total number of correct word pairs entered across the three trials (i.e., 12 x 3 = 36, a perfect score).

### MindCrowd: demographic, medical, health, and lifestyle questions

Upon completing the PAL task, participants were asked to fill out an additional 17 demographic and health/disease risk factor questions. These questions included: marital status, handedness, race, ethnicity, number of daily prescription medications, a first-degree family history of dementia, and yes/no responses to the following: seizures, dizzy spells, loss of consciousness (more than 10 minutes), high blood pressure, smoking status, diabetes mellitus, heart disease, cancer, reported stroke, alcohol/drug abuse, brain disease, and memory problems). Next, participants were shown their results and provided different comparisons to other test takers based on the average scores across all participants’ sex, age, and education demographics. On this same page of the site, participants were given the option to be recontacted for future research (see Supplementary Table 2 for the list of MindCrowd questions asked).

**Table 2.** MindCrowd Cohort Sample Sizes (n) for Each Factor. Summary of sample size numbers (*n*) of each multiple regression coefficients from MindCrowd.

| **Factor** | ***n*** |
| --- | --- |
| Biological Sex | Women = 47,700<br>Men = 27,966 |
| Educational Attainment | No High School Diploma = 1,881<br>High School Diploma = 6,695<br>Some College = 22,950<br>College Degree = 44,140 |
| Handedness | Left-Handed = 8,449<br>Right-Handed = 66,903 |
| Daily Medications Taken | None = 33,672<br>One = 14,409<br>Two = 9,651<br>Three = 6,656<br>Four = 10,769 |
| Reported Dizziness | Dizziness Reported = 4,749<br>No Dizziness Reported = 70,917 |
| Smoking Status | Smoker = 5,793<br>Non-Smoker = 69,873 |
| Diabetes Mellitus | Diabetes Reported = 3,887<br>No Diabetes Reported = 71,779 |
| Reported Stroke | Stroke Reported = 765<br>No Stroke Reported = 74,901 |

### UK Biobank: study design and aims

The UK Biobank (research ethics committee approval [11/NW/0382]) is a long-term study and research resource in the United Kingdom (UK), which investigates genetic and environmental exposure to the development of the disease. The UK Biobank’s stated goal is to “build a major resource that can support a diverse range of research intended to improve the prevention, diagnosis, and treatment of illness and the promotion of health throughout society.” The UK Biobank began in 2006. The study is currently following about 500,000 participants in the UK, enrolled at ages 40 to 69. Initial enrollment took place from 2006 to 2010. All participants are monitored for at least 30 years after recruitment and initial assessment (i.e., termed “instance 0” by the Biobank). Potential participants were invited to visit an assessment center, where they completed a questionnaire. Participants were next interviewed about lifestyle, medical history, and nutritional habits. Lastly, vital measurements, such as weight, height, and blood pressure, were measured. The UK Biobank aims to electronically record all health-related changes and events across the entire 30-year study. Notably, this task is aided by the UK’s integrated health system and corresponding electronic health record-keeping, an approach that is not possible in the United States.

### UK Biobank: data procurement

All UK Biobank data were derived from Application #43036, entitled “Exploring and Accommodating Heterogeneity in Large-Scale Genetic Analyses” as a “Collaborator Project,”

### UK Biobank: Complex visual recognition reaction time (cvrRT) and educational attainment

Each participant’s cvrRT was based on 12 rounds of the card-game’ Snap.’ Participants were shown two cards at a time with a picture on them. Participants pressed a button on a table in front of them as quickly as possible if the images cards/matched. For each of the 12 rounds, the following data were collected: the pictures shown on the cards (Index of card A, Index of card B), the number of times the participant clicked the ‘snap’ button, and the latency to first click of the ‘snap’ button. This last record of “latency to click the button” was used as the UK Biobank’s criterion for regression analyses.

For Educational Attainment, the following conversions from UK Biobank (UKBb) answer codes (see http://biobank.ndph.ox.ac.uk/showcase/coding.cgi?id=100305) to MindCrowd (MC) values were made: a) “UKBb **-7** None of the above” to “MC No high school diploma,” b) “UKBb **2** A levels/AS levels or equivalent” to “MC High school diploma,” c) “UKBb **3** O levels/GCSEs or equivalent” to “MC High school diploma,” d) “UKBb **4** CSEs or equivalent” to “MC High school diploma,” e) “UKBb **5** NVQ or HND or HNC or equivalent” to “MC Some college,” f) “UKBb **6** Other professional qualifications (e.g., nursing and teaching)” to” MC Some college,” g) “UKBb **1** College or University degree” to “MC College degree.” All UKBb participants selecting “**-3** Prefer not to answer” were removed from the final dataset before model selection and analysis. While we did our best to ensure a similar education measure across UKBb MindCrowd and the UK Biobank, we realize that there are fundamental differences between US and UK schools that we cannot control or eliminate. Table 6 lists the UK Biobank data fields used.

**Table 6.** UK Biobank Factor Generation. List of derived factors from the UK Biobank. UK Biobank’s “Index Number,” “Field Description,” “Field ID” and sample size (*n*s) for data field used to generate factors evaluated via multiple linear regression. “Data fields” were chosen to match to the questions posed in MindCrowd. If multiple instances (e.g., 0-4, timepoints) were available for data, only the first (instance 0) was used. For cvrRT and FHAD, multiple columns were compiled to generate the factor. For the UKBb MindCrowd and UK Biobank analyses, participants were removed if any of their responses to a demographic, medical, health, and lifestyle question did not match the other cohort.

| Index # | Field Description | Field ID | Count (n) | Type | Description | Column Label | Used to Generate |
| --- | --- | --- | --- | --- | --- | --- | --- |
| 6 | http://biobank.ndph.ox.ac.uk/showcase/field.cgi?id=31 | 31 | 502,524 | Categorical (single) | Sex | Sex | Biological Sex |
| 7 | http://biobank.ndph.ox.ac.uk/showcase/field.cgi?id=34 | 34 | 502,524 | Integer | Year of birth | Birth Year | Age |
| 264 | http://biobank.ndph.ox.ac.uk/showcase/field.cgi?id=404 | 404 | 202,464 | Integer | Duration to the first press of snap-button in each round | Snap-Button Time to First Press (Instance 0) | Median cvrRT (8 out 12 Rounds) |
| 395 | http://biobank.ndph.ox.ac.uk/showcase/field.cgi?id=1707 | 1707 | 501,626 | Categorical (single) | Handedness (chirality/laterality) | Handedness | Handedness |
| 422 | http://biobank.ndph.ox.ac.uk/showcase/field.cgi?id=2443 | 2443 | 501,593 | Categorical (single) | Diabetes diagnosed by doctor | Diabetes mellitus | Diabetes mellitus |
| 426 | http://biobank.ndph.ox.ac.uk/showcase/field.cgi?id=2453 | 2453 | 501,593 | Categorical (single) | Cancer diagnosed by doctor | Cancer | Cancer |
| 482 | http://biobank.ndph.ox.ac.uk/showcase/field.cgi?id=2966 | 2966 | 134,645 | Integer | Age high blood pressure diagnosed | Age Hypertension Diagnosed | Hypertension Diag. via MD |
| 616 | http://biobank.ndph.ox.ac.uk/showcase/field.cgi?id=4056 | 4056 | 7,579 | Integer | Age Reported Stroke diagnosed | Age Reported Stroke Diagnosed | Reported Stroke Diag. via MD |
| 2104 | http://biobank.ndph.ox.ac.uk/showcase/field.cgi?id=20107 | 20107 | 487,790 | Categorical (single) | Illnesses of father | (Multiple Columns) | FHAD |
| 2144 | http://biobank.ndph.ox.ac.uk/showcase/field.cgi?id=20110 | 20110 | 492,928 | Categorical (single) | Illnesses of mother | (Multiple Columns) | FHAD |
| 2188 | http://biobank.ndph.ox.ac.uk/showcase/field.cgi?id=20111 | 20111 | 433,922 | Categorical (single) | Illnesses of siblings | (Multiple Columns) | FHAD |
| 2325 | http://biobank.ndph.ox.ac.uk/showcase/field.cgi?id=20116 | 20116 | 501,633 | Categorical (single) | Smoking status | Smoking status | Smoking Status |
| 2356 | http://biobank.ndph.ox.ac.uk/showcase/field.cgi?id=21022 | 21022 | 502,524 | Integer | Age at recruitment | Age at Recruitment | Age (verify) |
| 120 | http://biobank.ndph.ox.ac.uk/showcase/field.cgi?id=401 | 401 | 495,702 | Categorical (single) | Index for card A in round | Needed to Know if Snap-Button Should Have Been Pressed | Median cvrRT |
| 168 | http://biobank.ndph.ox.ac.uk/showcase/field.cgi?id=402 | 402 | 495,702 | Categorical (single) | Index for card B in round | Needed to Know if Snap-Button Should Have Been Pressed | Median cvrRT |
| 1635 | http://biobank.ndph.ox.ac.uk/showcase/field.cgi?id=6138 | 6138 | 497,883 | Categorical (multiple) | Qualifications | Education | Educational Attainment |
| 2357 | http://biobank.ndph.ox.ac.uk/showcase/field.cgi?id=21053 | 21053 | 174,774 | Categorical (single) | Degree bothered by dizziness in the last three months | Dizziness | Dizziness |

### Data Quality Control

For the MindCrowd analysis, a final data set, including all qualifying participants up to 3-17-2020, was generated. See Supplementary Figure 9 for a flowchart detailing the following filtering steps. This dataset removed participants: a) with duplicate email addresses (only first entry kept), b) who did not complete all three rounds of the PAL test, c) whose primary language was not English, d) who was not between 18-85 years old, e) whose RT trials were above or below 1.5 x the interquartile range (IQR) and f) whose median svRT was above or below 1.5 x the IQR range of all participants of the same age (Supplementary Figure 10 details RT and IQR exclusion). Participants from either study were removed if they were missing any data (listwise deletion). Lastly, for the UKBb MindCrowd and UK Biobank analysis, participants were removed if their responses to a demographic, medical, health, and lifestyle question did not match the other study. For example, participants in the UK Biobank who responded to the “Race” question with “Prefer Not to Answer” were removed. “Prefer Not to Answer” was not a choice MindCrowd participants were given on the “Race” question. Removing these participants was done to align UKBb MindCrowd and UK Biobank cohorts as much as possible.

**Figure 9.**
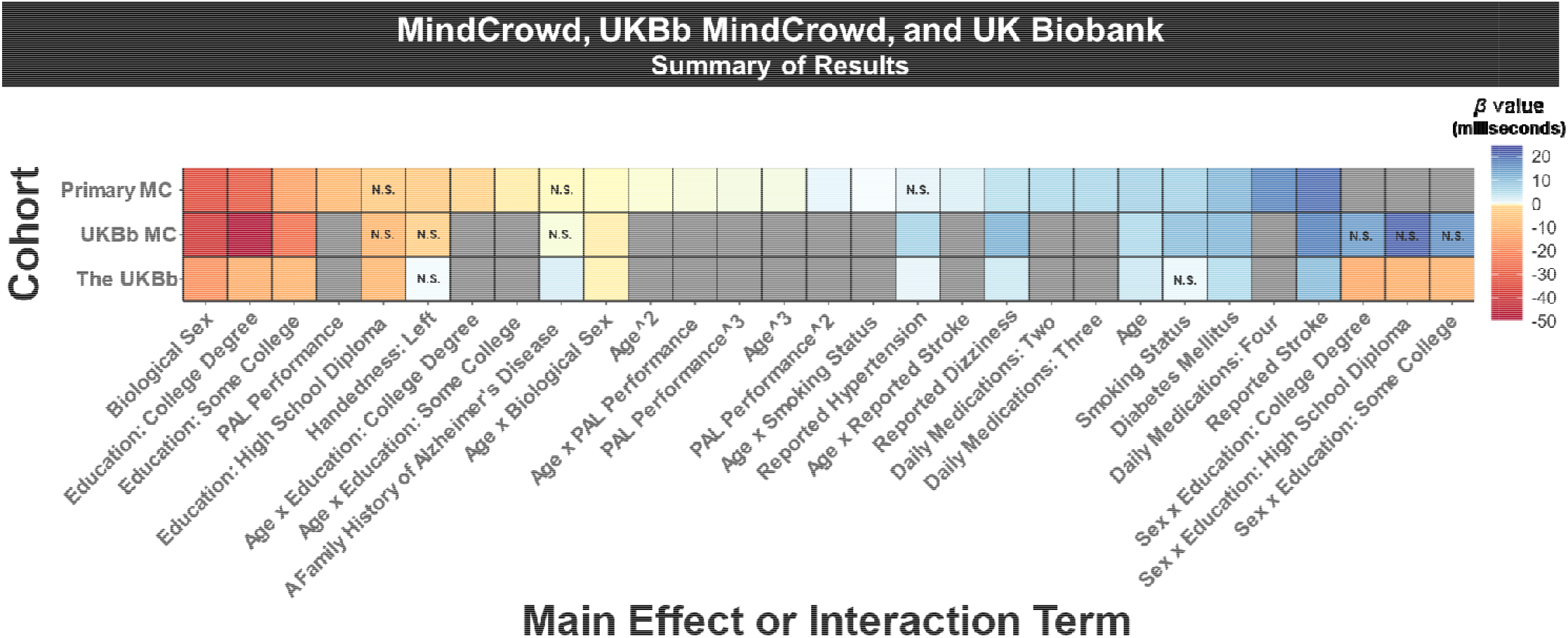
An illustrative summary of the results. Data are shown across the MindCrowd (MC), UKBbMindCrowd, and the UK Biobank (UKBb). The color (i.e., red = low negative and blue = high positive)indicates the size of the β (beta coefficient) estimate.” N.S.,” indicates if the estimated β value was not statistically significant (α = 0.05).

### Statistical Methods

Statistical analyses were conducted using R^36,37^. For all analyses, multivariate linear regression was performed using the general linear model (LM) to model Median svRT or Median cvrRT (i.e., criterion or dependent variable) as a function of either 24 (MindCrowd) or 11 predictors (UK Biobank analysis). For the MindCrowd analysis, svRT was modeled as a function of PAL Performance and Age raised to the power of three (i.e., to fit and estimate nonlinear associations). Most figures were created using “*ggplot2*” bundled together as a part of the R package, “*tidyverse*^38^”. Continuous by continuous interactions (i.e., simple slopes) were estimated using the R packages “*interactions*^*39*^,” “*sandwich*^*40*^,” “*jtools*^41^". Categorical by categorical interactions were estimated using the R package, “*emmeans*” ^42^. Adjustments for multiple comparisons were evaluated using Tukey’s method via the “*emmeans*” package. Missing data were assessed via the “*finalfit*,^43^” “*visdat*,^*44*^” and “*naniar*^45^” R packages (see Supplementary Table 1 for a complete list of resources).

**Table 1.** Sociodemographic Characteristics of Each Cohort. Sociodemographic Characteristics Across Each of the Three Cohorts. Study *n*s and related percentages of commonly reported sociodemographic factors from each of the three cohorts (i.e., MindCrowd and UKBb MindCrowd and the UK Biobank).

| *Cohort* | *Descriptive or Factor Level* | *n* | *%* |
| --- | --- | --- | --- |
| **1. MindCrowd 18-85yr: Age** | *M* = 46.54 *SD* = 18.40 | 75,666 | 100 |
| UKBb MindCrowd 40-70yr: Age | *M* = 56.59 *SD* = 8.16 | 39,795 | 100 |
| UK Biobank 40-70yr: Age | *M* = 55.81 *SD* = 7.72 | 158,249 | 100 |
| **2. MindCrowd 18-85yr: Biological Sex** | Women | 47,700 | 63.08 |
|  | Men | 27,966 | 36.91 |
| UKBb MindCrowd 40-70yr: Biological Sex | Women | 29,640 | 74.51 |
|  | Men | 10,155 | 25.49 |
| UK Biobank 40-70yr: Biological Sex | Women | 89,333 | 56.45 |
|  | Men | 68,916 | 43.55 |
| **3. MindCrowd 18-85yr: Race** | Native American | 445 | 0.6 |
|  | Asian | 3,491 | 4.74 |
|  | Black/African American | 1,555 | 2.11 |
|  | Native Hawaiian/Pacific Islander | 270 | 0.37 |
|  | Mixed | 490 | 0.66 |
|  | White | 67,433 | 91.52 |
| UKBb MindCrowd 40-70yr: Race | Asian | 743 | 1.90 |
|  | Black/African American | 732 | 1.87 |
|  | Mixed | 182 | 0.47 |
|  | White | 37,235 | 95.03 |
| UK Biobank 40-70yr: Race | Asian | 1,612 | 1.02 |
|  | Black/African American | 979 | 0.62 |
|  | Mixed | 847 | 0.54 |
|  | White | 154,809 | 97.83 |
| **4. MindCrowd 18-85yr: FHAD** | TRUE | 17,382 | 23.59 |
|  | FALSE | 56,302 | 76.41 |
| UKBB MindCrowd 40-70yr: FHAD | TRUE | 13,554 | 34.59 |
|  | FALSE | 25,627 | 65.41 |
| UK Biobank 40-70yr: FHAD | TRUE | 19,741 | 12.48 |
|  | FALSE | 138,506 | 87.52 |
| **5. MindCrowd 18-85yr: Handedness** | Left | 8,276 | 11.23 |
|  | Right | 65,408 | 88.77 |
| UKBb MindCrowd 40-70yr: Handedness | Left | 4,474 | 11.42 |
|  | Right | 34,707 | 88.58 |
| UK Biobank 40-70yr: Handedness | Left | 15,287 | 9.66 |
|  | Right | 142,960 | 90.34 |
| **6. MindCrowd 18-85yr: Educational Attainment** | No High School Diploma | 1,792 | 2.43 |
|  | High School Diploma | 6,470 | 8.78 |
|  | Some College | 22,335 | 30.31 |
|  | College Degree | 43,087 | 58.48 |
| UKBb MindCrowd 40-70yr: Educational Attainment | No High School Diploma | 577 | 1.47 |
|  | High School Diploma | 3,108 | 7.93 |
|  | Some College | 10,955 | 27.96 |
|  | College Degree | 24,541 | 62.63 |
| UK Biobank 40-70yr: Educational Attainment | No High School Diploma | 10,979 | 6.94 |
|  | High School Diploma | 46,247 | 29.23 |
|  | Some College | 77,271 | 48.83 |
|  | College Degree | 23,750 | 15.01 |

### Model evaluation and controls

All measurements were taken from distinct samples. Model fit and violations of parametric assumptions were evaluated separately in each model. Here we evaluated different residual plots, assessing normality, homoscedasticity, outliers, residual autocorrelation, and multicollinearity. The MindCrowd LM included all 22 demographic questions, health, medical, and lifestyle questions. These questions were: Age, Biological Sex, Race, Ethnicity, Educational Attainment, Marital Status, Handedness, Number of Daily Medications, Seizures, Reported Dizziness, Loss of Consciousness, Reported Hypertension, Smoking Status, Heart Disease, Reported Stroke, Alcohol/Drug Abuse, Diabetes Mellitus, Cancer, a First-Degree Family History of Alzheimer’s disease, history of brain disease, whether the test was taken on a mobile device, and the version of the MindCrowd site used. Not surprisingly, the device used to take the RT test in MindCrowd was associated with RT performance. For the UK Biobank analyses, these 11 variables included: Age, Biological Sex, Diabetes mellitus, Handedness, Reported Stroke, Reported Hypertension, Smoking Status, Reported Dizziness, Educational Attainment, and a First-Degree Family History of Alzheimer’s Disease. Examination of each model’s variance inflation factors (VIF) revealed no unexpected factors with a VIF > 5 (i.e., considered “highly” colinear by convention, see Supplementary Table 3).

**Table 3.**
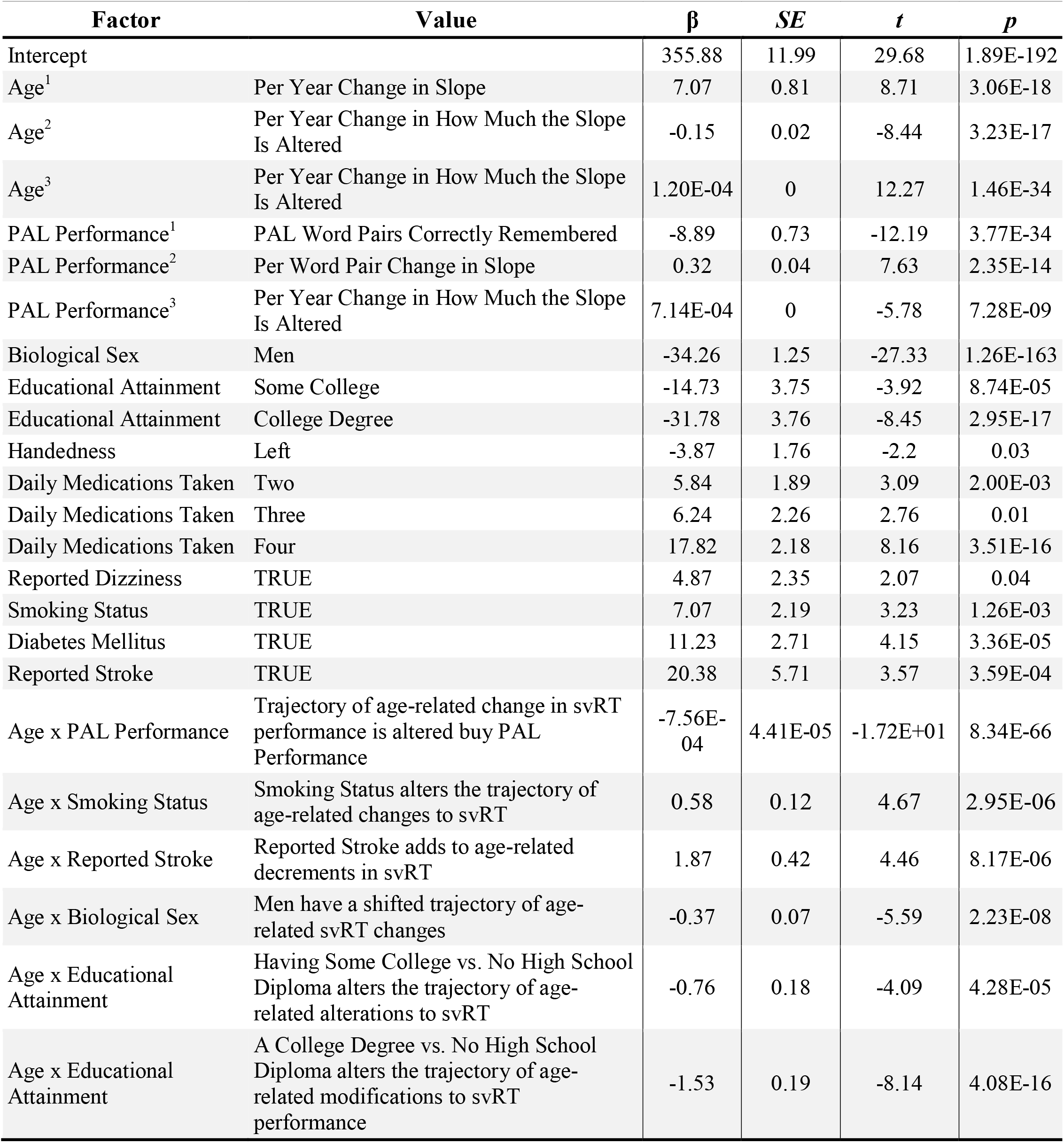
MindCrowd Summary of Results. Summary of the main findings from MindCrowd, listing main associations and interactions. β = unstandardized regression coefficient, *SE* = standard error, *t* = value of *t*-statistic, *p* = *p*-value.

### Inclusion of polynomials and automated model selection

The MindCrowd analyses included Age and PAL Performance as first through third-degree non-orthogonal polynomials (i.e., cubic regression). This choice was based on empirical evaluations, using Bayesian information criterion (BIC) weights (a.k.a Schwarz weights)^46^. We generated, ran, and recorded BICs across seven models (i.e., base and first – sixth-degree-[nonorthogonal]polynomial). BIC weights were calculated from raw BIC values using the “*qPCR*^*47*^” (v1.4-1) R for each model. The third-degree polynomial model reported the largest BIC weight, and it was 1.46E+270 times more likely to occur than the base (no polynomial) model (_BICHighW_ 9.99E-01 / _BICLowW_6.82E-271 = 1.46E+270) ^46^. It is worth noting that a prior study examining both complex and simple RT also included age as a third-degree polynomial. Other similarities included: a relatively large *n* = 7,000, both sexes, an 18-94 years-old age-range, and several RT findings^46^.

For the MindCrowd, UKBb MindCrowd, and UK Biobank cohorts, we used the R package “glmulti” (v1.0.8)^48^ to define our GLM models. *“glmulti”* uses full information criterion model selection versus shrinkage regression methods (e.g., LASSO or LAR)^48^. We used *“glmulti”* to avoid the pitfalls of stepwise selection methods or unintentional biased introduced via manual or *p*-value-based model selection. We had *“glmulti”* define the “best” (i.e., lowest *BIC*) MindCrowd and UK Biobank models separately using its *“genetic”* algorithm method with *“marginality”* set to True. We chose *BIC* as opposed to other information criterion methods because *BIC* punishes for model complexity. Two rounds of model selection were run to find pairwise interactions due to package limitations (i.e., millions of potential models). For round 1, the optimal model contained only the main effects when all 22 factors were included. In round 2, the only factors selected in the optimal main effects model were then included to select an optimal model, including two-way interactions.

### Code availability

The code supporting each analysis, figure, table, or other material is publicly available. The directory containing these files is included in this study’s data repository (see Data availability for more information).

## Results

### MindCrowd

As of March 13th, 2020, after filtering (see Data Quality Control in Materials and Methods), MindCrowd, had recruited 75,666 qualified participants (see Table 1 for Sociodemographic Characteristics and Supplementary Figure 1A for a histogram of age). We modeled svRT as a function of Age^3^ and PAL Performance^3^ (i.e., curvilinear associations) as well as 20 other factors (see Supplementary Figure 2 for diagnostic regression plots, and Table 2 for all coefficient *n*s). The omnibus model was significant (*F*_*omnibus*_[58, 73406] = 858.20, *p*_*omnibus*_ < 2.2e-16, Adjusted *R*^2^ = 0.40).

**Figure 1.**
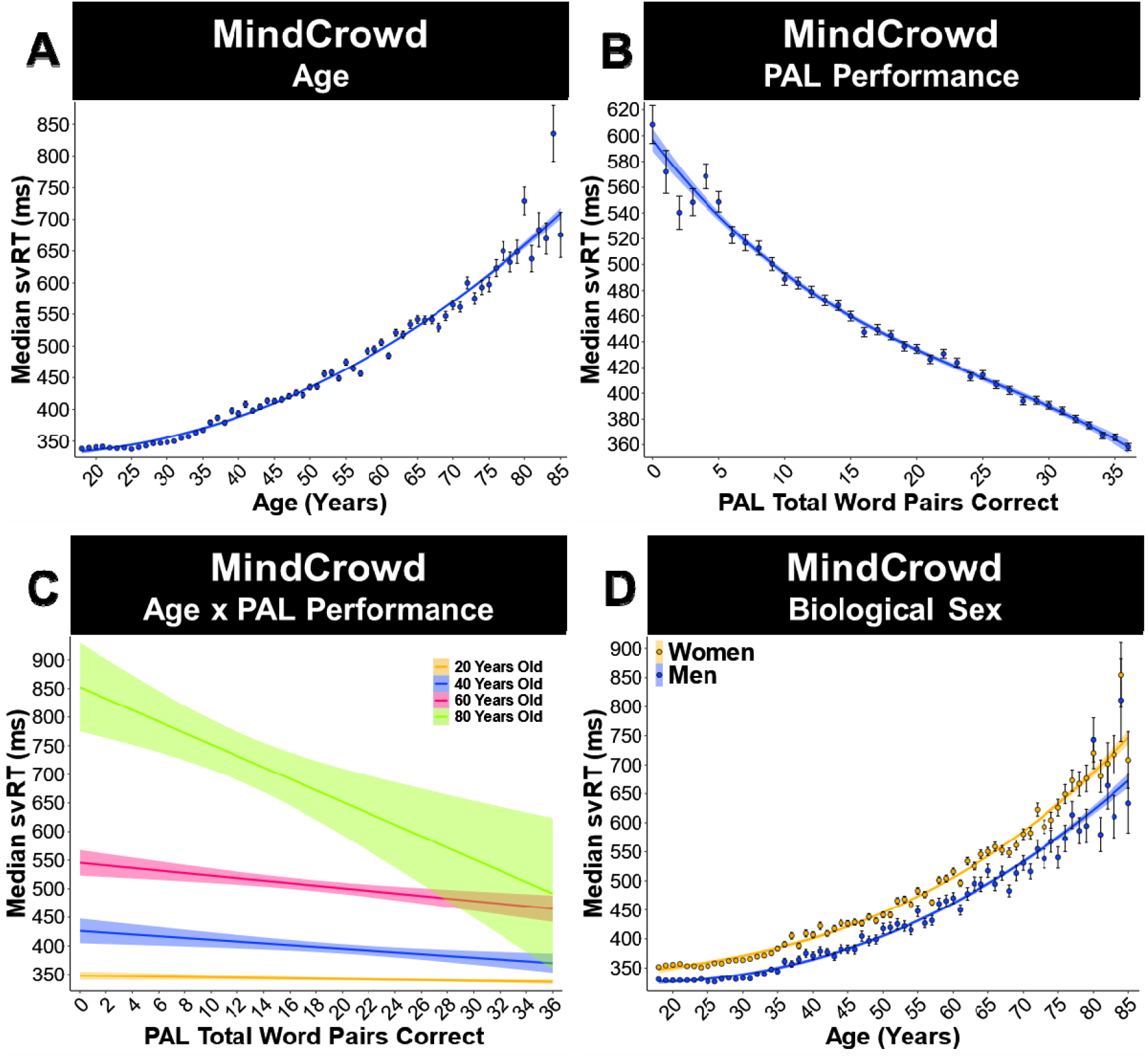
Age, paired associates learning (PAL) performance, and Biological Sex were associated with simple visual reaction time (svRT) in the MindCrowd analysis (ages 18-85). (A) Linear model fits (line fill ± 95% CI, error bars ± SEM) of the median svRT by Age3 (curvilinear model). There was a curvilinear relationship between svRT and Age^1^ (β_Age_1 = 7.07, *p*_Age_1 = 3.06E-18), Age^2^ (β_Age2_ = -0.15, p_Age2_ = 3.23E-17), and Age^3^ (β_Age3_ = 1.47E-03, *p*_Age3_< 1.46E-34, *n* = 75,666). (B) Linear model fits (line fill ± 95% CI, error bars ± EM) ofthe median svRT by Age3 (curvilinear model). There was a curvilinear relationship between svRT and paired-associate learning (PAL) performance PAL Performance^1^ (β_PAL1_ = -8.89, *p*_PAL1_ = 3.77E-34), PAL Performance^2^ (β_PAL2_ = 0.32, *p*_PALl2_ = 2.35E-14), and PAL Performance^3^ (β_PAL3_ = -4.13E-03, *p*_PAL3_ = 7.28E-09, *n* = 75,666). (C)Simple slope analysis of the Linear model fits (line fill ± 95% CI, error bars ± SEM) of the median svRT by PAL Performance (β_Age*PAL_ = -0.07, *p*_Age*PAL_ = 1.26E-59). At 20 (β_Age20*PAL_ = -5.48, *p*_Age20*PAL_ < 0.00, *n* = 1985), 40 (β_Age40*PAL_ = -6.41, *p*_Age40*PAL_ < 0.00, *n* = 739), 60 (β_Age60*PAL_ = -7.75, *p*_Age60*PAL_ < 0.00, *n* = 1789), and80 (β_Age80*PAL_ = -9.08, *p*_Age80*PAL_ < 0.00, n = 344) years of age. (D) Linear model fits (line fill ± 95% CI, errorbars ± SEM) of the median svRT by Age^3^ (curvilinear model) with lines split by Biological Sex. Womendemonstrated longer svRT compared to men from younger to older ages (β_Sex_ = -34.26, *p*_Sex_ = 1.26E-163, *n*_Women_= 47,700, *n*_Men_ = 27,966).

**Figure 2.**
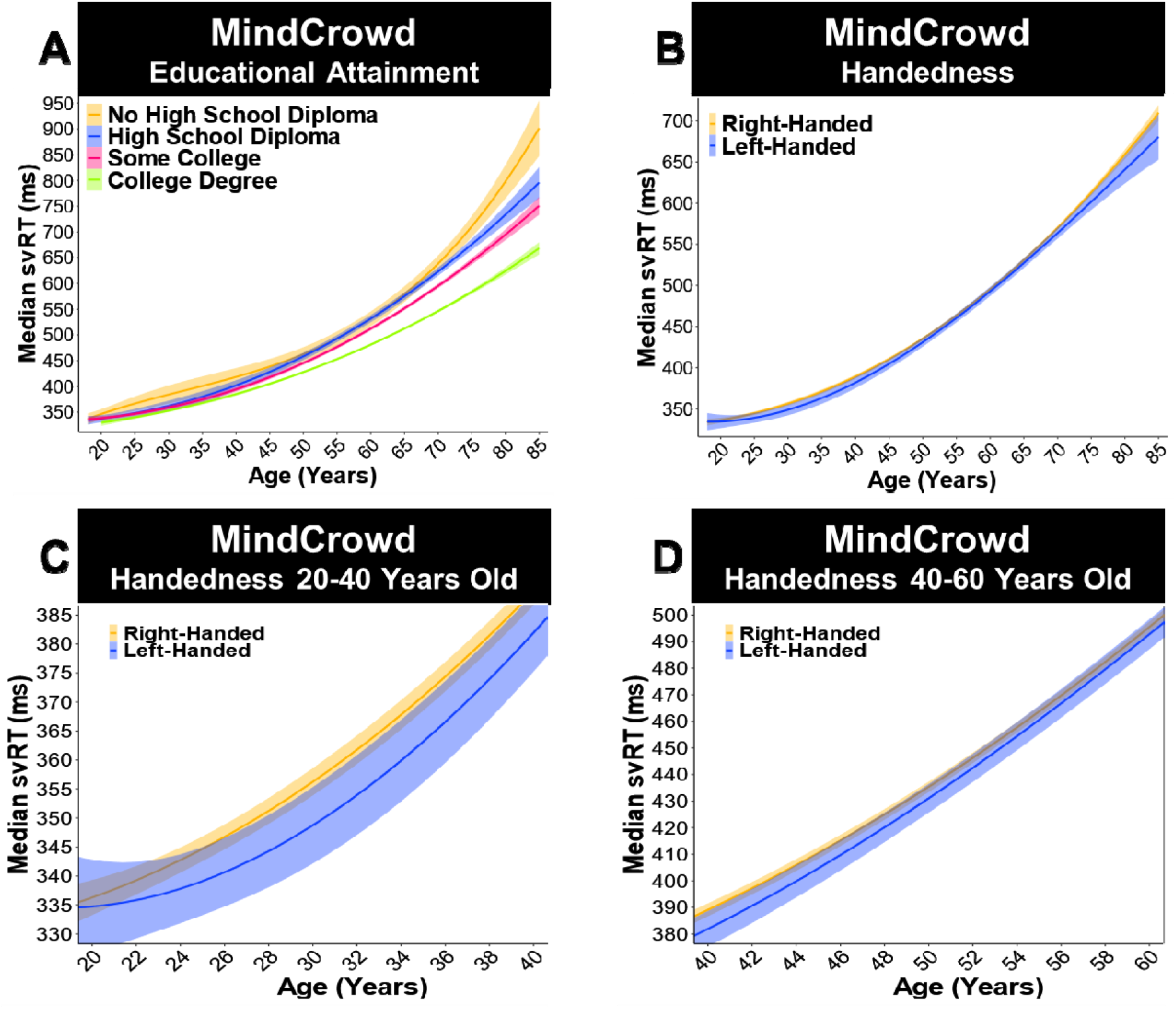
Educational attainment and handedness altered simple visual reaction time (svRT) performance. MindCrowd analysis (ages 18-85). (A) Linear model fits (line fill ± 95% CI) of the median svRT by Age^3^ (curvilinear model) with lines split by Educational Attainment. Participants who had “Some College” (β_College_ =-14.73, *<u>p</u>*_College_ = 8.74E-05, *n* = 22,950), or a “College Degree” (β_CDegree_ = -31.78, *p*_CDegree_ = 2.95E-17, *n* = 44,140) were faster than those with “No High School Diploma” (*n* = 1,881). (B-D) Linear model fits (line fill ± 95% CI) of the median svRT by Age^3^ (curvilinear model) with lines split by Handedness. (B) From 18-85 years old, left-handed participants showed slightly faster svRTs (β_Left_ = -3.87, *p*_Left_ < 0.03), (C) an association found in 20-40 years olds (β_40Left_ = -3.16, *p*_40Left_ < 0.01, Left-Handed *n* = 8,449), (D) but not 40-60 years olds (β_60Left_ =-2.69, *p*_60Left_ = 0.07, Right-Handed *n* = 66,903).

### Curvilinear associations: Age and paired-associate learning (PAL) total correct

Our model revealed that all three Age polynomials were significantly associated with svRT. Age^1^ (i.e., linear association, first-degree polynomial, aka slope), Age^2^ (i.e., quadratic association, second-degree polynomial), and Age^3^ (i.e., cubic association, third-degree polynomial). On average from younger to older Age, a one-year difference (X=1). (1) Age^1^ (shift in Y; *p*_*Age2*_ = 3.23E-17) was associated with 7 msec longer svRT. (2) Age^2^ (shift in Age^1^; *p*_*Age1*_ = 3.06E-18) was associated with 0.15 msec of additional svRT length (i.e., 7 + .15 msec/year, Figure 1A). (3) Age^3^ (shift in Age^2^; *p*_*Age3*_ = 1.46E-34) was associated with a negligible 1.20E-04 msec shift in additional svRT length (i.e., 7 + (0.15 + 1.20E-4) msec/year, Figure 1A). In contrast to Age’s association with longer svRT, each word pair correct for PAL Performance. (1) PAL^1^ (*p*_*PAL1*_ = 3.77E-34) was associated 9 msec shorter svRT. (2) PAL^2^ (*p*_*PAL2*_ = 2.35E-14) was associated with 0.32 msec of additional svRT shortening (i.e., 9 + 0.32 msec/year, Figure 1B). (3) PAL^3^ (*p*_*PAL3*_ = 7.28E-09) was associated with a slight 7.14E-04 msec shift in additional svRT shortening (i.e., 9 + (0.32 + 7.14E-04) msec/year Figure 1B).

### Biological sex, educational attainment, and handedness

Biological Sex was a significant predictor of svRT (*p*_*Sex*_ = 1.26E-163). Being a man was associated with an average of 34 msec (9.63%) faster svRT response than being a woman (Figure 1D). Educational Attainment was also a significant factor associated with svRT. Compared to “No High School Diploma,” participants who had “Some College” (*p*_*College*_ = 8.74E-05), or a “College Degree” (*p*_*CDegree*_ = 2.95E-17, Figure 2A) were faster. Attending college and attaining a college degree was associated with a respective near 15 (4.14%) and 32 (8.92%) msec shorter svRT compared to not graduating from high school. Handedness was also associated with svRT. Left-handed participants had a near 4 msec (1.09%) faster svRT (*p*_*Left*_ = 0.03, Figure 2B). This association was present in individuals 20 to 40 years old (*p*_*40Left*_ < 0.01, Figure 2C) but not in individuals 40 to 60 years old (*p*_*60Left*_ = 0.07, Figure 2D).

### Health, medical, and lifestyle factors

For health and medical factors associated with svRT, we found that Smoking Status (*p*_*Smoking*_ = 1.26E-03, Figure 3A) and Reported Dizziness (*p*_*Dizzy*_ = 0.04, Figure 3B) were both significant predictors of svRT. Smoking Status was associated with 7 msec (1.99%) slower svRT, and Reported Dizziness was associated with nearly 5 msec (1.37%) slower svRT. When compared to participants reporting “no daily medications,” taking “Two” (*p*_*Meds2*_ = 2.00E-03), “Three” (*p*_*Meds3*_ < 0.01), and “Four” (*p*_*Meds4*_ = 3.51E-16) Daily Medications were associated with an approximately 5 (1.64%), 6 (1.76%), and 18 (5.01%) msec longer svRTs, respectively (Figure 3C). Further, Diabetes Mellitus (*p*_*Diabetes*_ < 3.36E-05, Figure 3D), and Stroke (*p*_*Stroke*_ = 3.59E-04, Figure 3E) were related to 11 (3.16%) and 20 (5.73%) msec longer svRTs respectively when compared to participants not reporting either condition. Of note, in this model, both a first-degree family history of Alzheimer’s disease (FHAD; *p*_*FHAD*_ = 0.78) and Hypertension (*p*_*Hyper*_ = 0.52) were not significant predictors of svRT performance (Supplementary Figure 3A-B).

**Figure 3.**
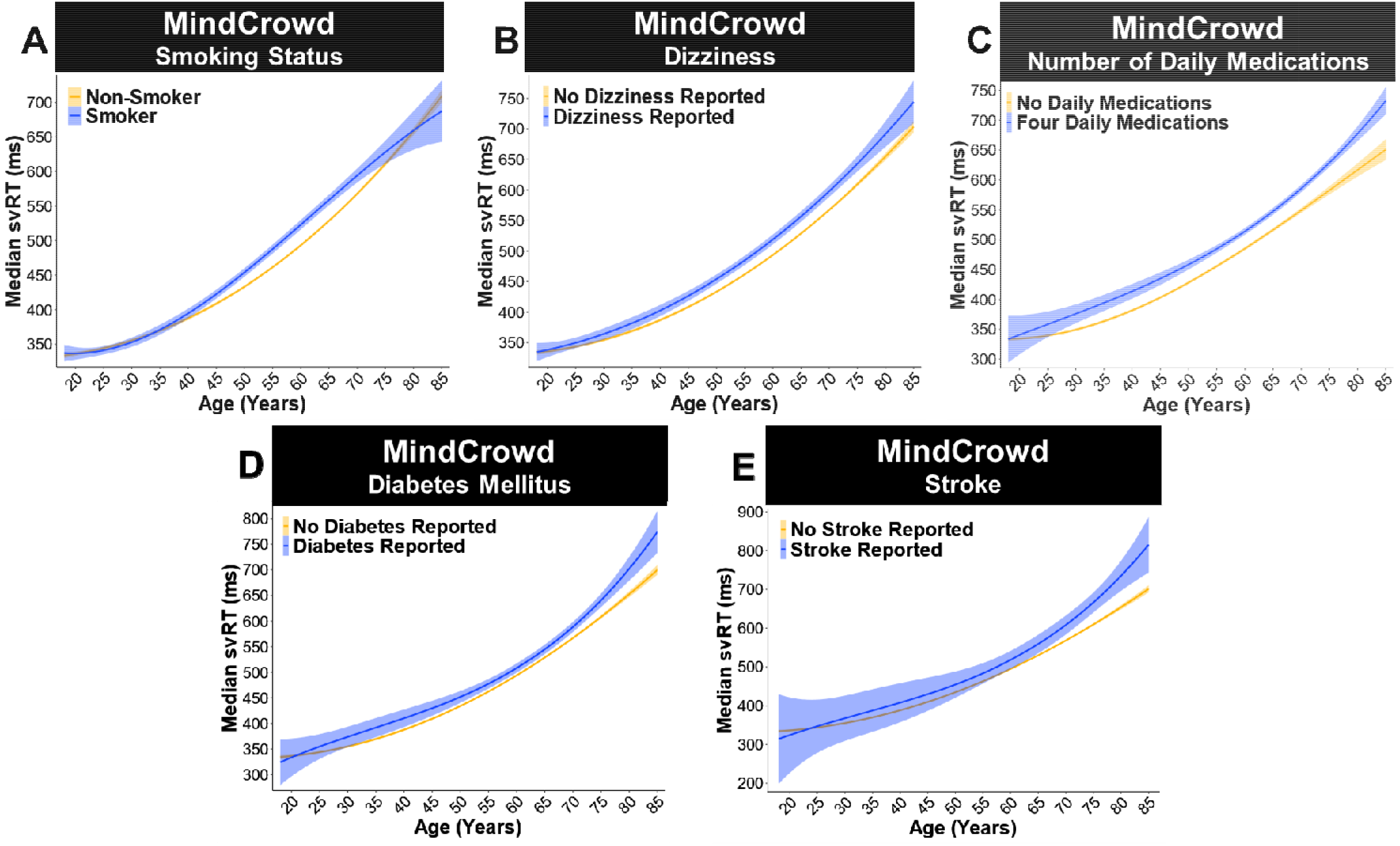
***A-B*** Simple visual reaction times (svRT) were slower for those who smoked or Reported Dizziness. MindCrowd analysis (ages 18-85). (A) Linear model fits (line fill ± 95% *CI*) of the median svRT by Age^3^ (curvilinear model) with lines split by Smoking Status. Participants identifying as a smoker showed slower svRT (β_*Smoking*_ = 7.07, *p*_*Smoking*_ = 1.26E-03, Smoker *n =* 5,793, Non-Smoker *n =* 69,873). (B) Linear model fits (line fill ± 95% *CI*) of the median svRT by Age^3^ (curvilinear model) with lines split by Reported Dizziness.Participants reporting Reported Dizziness showed slower svRT performance (β_*Dizzy*_ = 4.87, *p*_*Dizzy*_ = 0.04,Reported Dizziness Reported *n =* 4,749, No Reported Dizziness Reported *n =* 70,917). ***C-E*** Health factors suchas daily medications taken, diabetes, and Reported Stroke all slowed simple visual reaction time (svRT).MindCrowd analysis (ages 18-85). (C) Linear model fits (line fill ± 95% *CI*) of the median svRT by Age^3^ (curvilinear model) with lines split by Daily Medications Taken. Compared to participants reporting “no daily medications” (*n =* 33,672), taking “Two” (β_*Meds2*_ = 5.84, *p*_*Meds2*_ = 2.00E-03, *n =* 9,651), “Three” (β_*Meds3*_ = 6.24, *p*_*Meds3*_<0.01, *n =* 6,656), and “Four” (β_*Meds4*_ = 17.82, *p*_*Meds4*_ = 3.51E-16, *n =* 10,769) slowed svRT. (D) Linear model fits (line fill ± 95% *CI*) of the median svRT by Age^3^ (curvilinear model) with lines split by diabetes mellitus. Participants reporting having diabetes had slower svRT performance (β_*Diabetes*_ = 11.23, *p*_*Diabetes*_<3.36E-05, Diabetes Reported *n =* 3,887, No Diabetes Reported *n =* 71,779). (E) Linear model fits (line fill ± 95% *CI*) of the median svRT by Age^3^ (curvilinear model) with lines split by Reported Stroke. Reporting having had aReported Stroke impaired (slowed) svRT performance (β_*Stroke*_ = 20.38, *p*_*Stroke*_ = 3.59E-04, Reported Reported *n =* 765, No Reported Stroke Reported *n =* 74,901).

### Two-way interactions

For interactions, we found Age significantly interacted with PAL Performance. Age x PAL Performance (*p*_*Age*PAL*_ = 9.93E-62). Analysis of simple slopes suggests that each word-pair correct was associated with shorter svRT from younger to older ages. That is, at 20 (*p*_*Age20*PAL*_<0.00),40(*p*_*Age40*PAL*_<0.00),60(*p*_*Age60*PAL*_<0.00), and 80 (*p*_*Age80*PAL*_<0.00, Figure 1C) years old. There was a significant Biological Sex x Age interaction (*p*_*Age*Sex*_ = 4.61E-08), indicating that the associated slowing of svRT at younger and older ages in men, compared to women, was 0.36 msec (0.08%) less per one year difference in age (Figure 1C). These data suggest that men’s svRT lengthened at a slower rate when compared to women. Of interest, in both women and men, we found significant Age x Educational Attainment interactions. Compared to Age x “No High School Diploma", participants reporting having “Some College” (*p*_*Age*College*_ = 4.20E-04) or a College Degree” (*p*_*Age*CDegre*_ = 2.07E-12) was associated with slower RTs from young to an older age. These results suggest that attending college or getting a college degree was associated with a 0.65 (0.15%) and 1.31 (0.30%) msec faster svRT performance per year of life, respectively (Figure 2A). The MindCrowd model revealed a significant Age x Reported Stroke interaction (*p*_*Age*Stroke*_ = 2.72E-06). Participants who Reported Stroke were about 2 msec (0.37%) slower on svRT per year of life than those without Reported Stroke (Figure 3E). Lastly, we found a significant Age x Smoking Status interaction (*p*_*Age*Smoke*_ = 5.67E-07). This interaction suggests that Smoking Status lengthens svRT by adding 0.57 msec (0.11%) per year difference in age (Figure 3A). See Table 3 for a complete list of the significant predictors, covariates, and interactions from the MindCrowd analysis.

### Mobile Device

Participants who were identified as using a mobile device to take MindCrowd (i.e., using a touchscreen, *n* = 7,603, age *M* = 54.06 *SD* = 14.54 years) had slower svRTs and were older (β_Age∼Mobile_ = 14.13, *p*_Age∼Mobile_ < 2e-16) than those who did not use a mobile device (*n* = 76,775, age *M* = 45.54 *SD* = 18.43 years, see Supplementary Figure 8).

**Figure 8.**
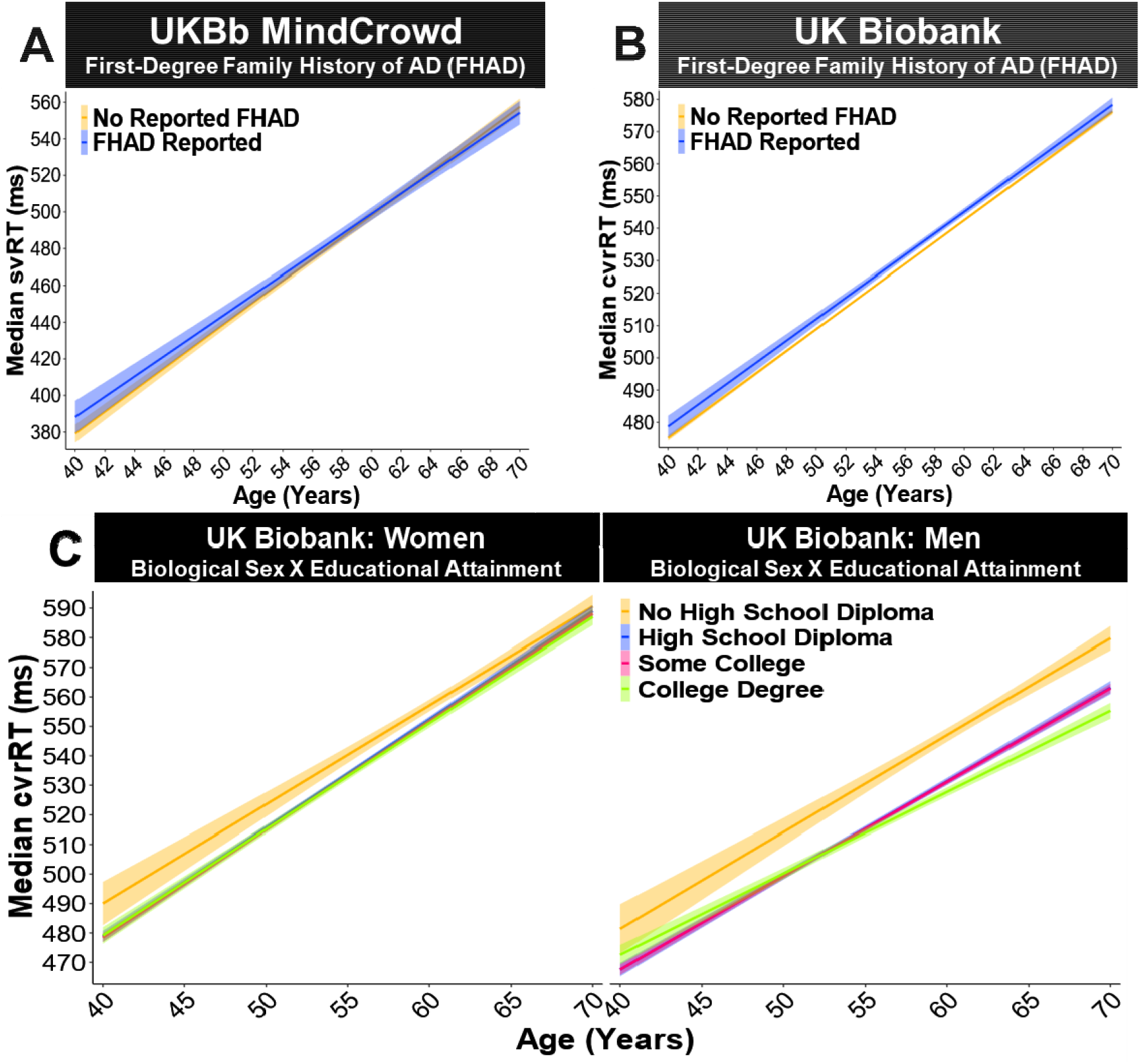
***A-B*** A first-degree family history of Alzheimer’s disease (FHAD) was related to slower UK B complex, visual recognition reaction time (cvrRT), UKBb MindCrowd simple visual reaction time (svR Biobank analysis (ages 40-70). (A) UKBb MindCrowd linear model fits (line fill ± 95% *CI*) of median svRT across Age with lines split by reported FHAD. An association between FHAD and svRT was not found in the UKBb MindCrowd cohort (β_*FHAD*_ = -0.07, *p*_*FHAD*_ = 0.97, FHAD Reported *n =* 13,748, No FHAD Reported *n =* 26,047). (B) UK Biobank linear model fits (line fill ± 95% *CI*) of median cvrRT across Age with lines split by reported FHAD. Compared to those reporting No FHAD, FHAD was related to worse cvrRT performance in the UK Biobank (β_*FHAD*_ = 2.36, *p*_*FHAD*_ = 3.35E-05, FHAD Reported *n =* 19,741, No FHAD Reported *n =* 138,504). ***C*** In the UK Biobank, biological sex modified the association of educational attainment and complex, visual, recognition reaction time (cvrRT) (ages 40-70). Linear model fits (line fill ± 95% *CI*) of the median cvrRT by Age with lines split by Educational Attainment and Biological Sex. Compared to women having “No High School Diploma” (*n =* 6,056), men with a “High School Diploma” (β_*Sex*HSDiploma*_ *=* -11.24, *p*_*Sex*HSDiploma*_ = 3.30E-12, *n =* 18,505), “Some College” (β_*sex*College*_ = -10.54, *p*_*sex*College*_ = 8.71E-12, *n =* 34,230) or a “College Degree” (β_*sex*CDegree*_ = -12.8, *p*_*sex*CDegre*_ = 1.68E-13, *n =* 11,257) were all significantly associated with faster cvrRT. See Supplementary Figure 7, displaying simple effects parsed using estimated marginal means (*EMM*).

### UKBb MindCrowd and UK Biobank

Of the total 75,666 MindCrowd participants, 39,759 between the ages of 40 and 70 were selected to mirror the UK Biobank. This subset is called UKBb MindCrowd from here on to differentiate it from MindCrowd. After filtering (see Data Quality Control), the UK Biobank cohort had 158,249 participants, derived from a data-request we received on 9-19-2019 (See Table 1 for Sociodemographic Characteristics and Supplementary Figure 1B-C for age histograms). We model both the UKBb MindCrowd’s svRT (see Supplementary Figure 4 for regression diagnostic plots) as well UK Biobank’s cvrRT (see Supplementary Figure 5 for regression diagnostic plots) as a function of 11 shared survey questions (see Table 4 for the *n*s used to estimate each coefficient). The omnibus UKBb MindCrowd (*F*_*mcomni*_[20, 38871] = 1039, *p*_*mcomni*_ < 2.2e-16, Adjusted *R*^2^ = 0.08) and UK Biobank (*F*_*ukbbbomni*_[20, 157903] = 1038, *p*_*ukbbomni*_ < 2.2e-16, Adjusted *R*^2^ = 0.13) LMs were both significant. Table 5 lists the effects for each cohort side by side.

**Table 4.**
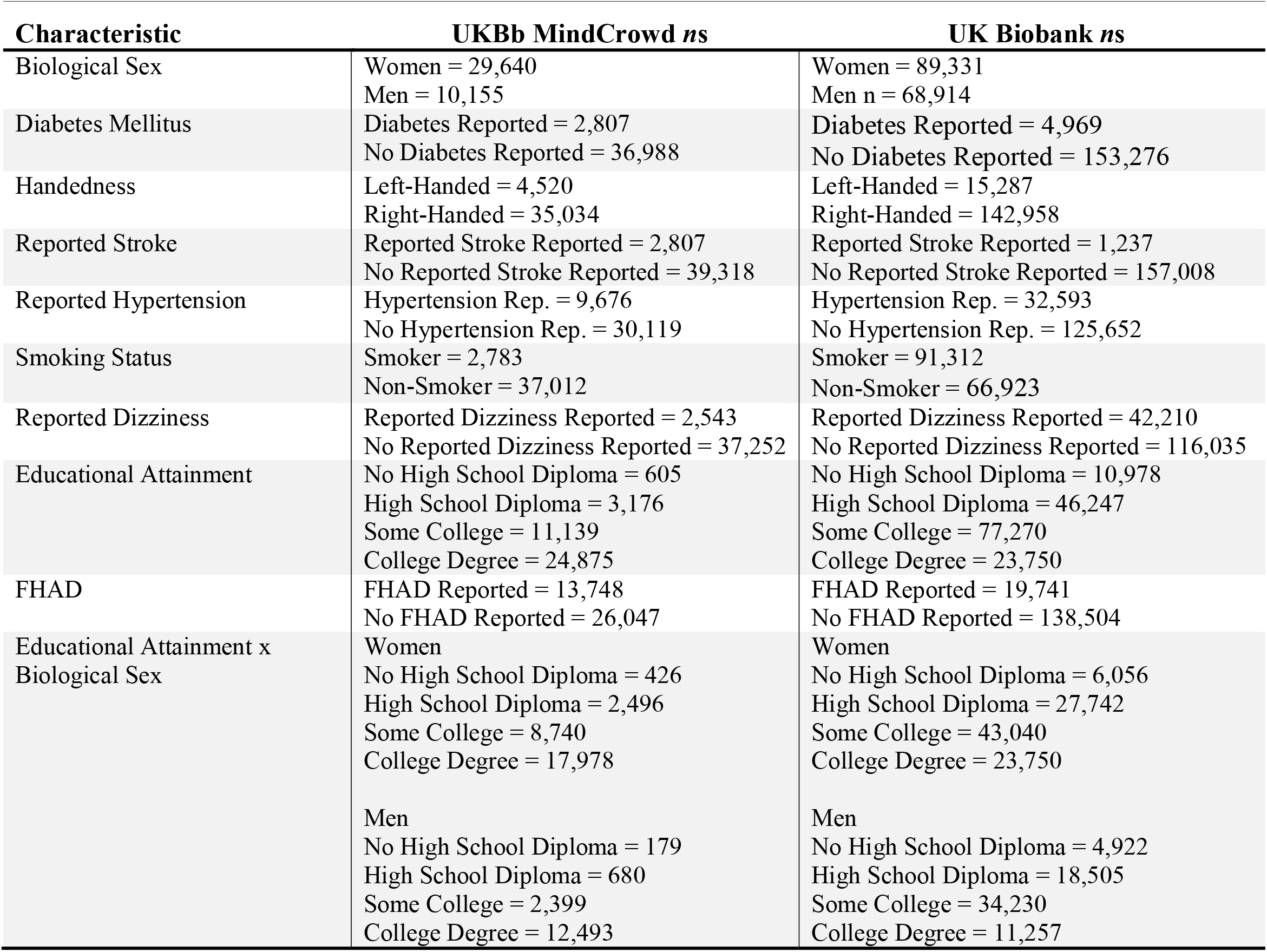
UKBb MindCrowd and UK Biobank Cohort Sample Sizes (n) Summary of sample size numbers (*n*s) of each multiple regression coefficient from the UKBb MindCrowd and UK Biobank.

**Table 5.** UKBb MindCrowd and the UK Biobank Summary of Results. Summary of the main findings from the UKBb MindCrowd and the UK Biobank cohorts. Here factor names, factor definitions, β = unstandardized regression coefficients as well as estimate *SE* = standard error, *t* = value of *t*-statistic, and *p* = *p*-value across both cohorts is shown. The last column displays model outcomes in terms of the agreement between the cohorts.

| Variable | Value | UKBb MindCrowd |  |  |  | The UK Biobank |  |  |  | Outcome |
| --- | --- | --- | --- | --- | --- | --- | --- | --- | --- | --- |
| | | $\beta$ | *SE* | *t* | *p* | $\beta$ | *SE* | *t* | *p* | |
| Intercept |  | 189.77 | 10.65 | 18.35 | 6.46E-75 | 335.93 | 1.69 | 209.93 | 2.00E-16 | Agree |
| Age | Per Year | 5.75 | 0.12 | 46.57 | 2.00E-16 | 3.40 | 0.03 | 132.87 | 2.00E-16 | Agree |
| Biological Sex | Men | -40.00 | 2.24 | -17.83 | 8.03E-71 | -18.28 | 0.38 | -47.70 | 2.00E-16 | Agree |
| Diabetes Mellitus | TRUE | 11.48 | 3.91 | 2.94 | 3.31E-03 | 5.48 | 1.09 | 5.03 | 4.80E-07 | Agree |
| Handedness | Left | -2.58 | 3.05 | -0.85 | 4.00E-01 | 0.58 | 0.63 | 0.92 | 0.36 | N.S. in both - but MindCrowd subtle effect |
| Reported Stroke | TRUE | 18.47 | 9.02 | 2.05 | 4.00E-02 | 10.61 | 2.13 | 4.99 | 6.15E-07 | Agree |
| Reported Hypertension | TRUE | 6.99 | 2.37 | 2.95 | 3.16E-03 | 1.09 | 0.48 | 2.27 | 2.00E-02 | Agree |
| Smoking Status | TRUE | 10.18 | 3.87 | 2.63 | 0.01 | 0.40 | 0.38 | 1.05 | 0.29 | MindCrowd Finding: Both Primary and UKBb |
| Reported Dizziness | TRUE | 12.10 | 4.00 | 3.02 | 2.52E-03 | 3.21 | 0.42 | 7.57 | 3.71E-14 | Agree |
| Educational Attainment | High School Diploma | -9.68 | 8.77 | -1.10 | 2.70E-01 | -9.68 | 0.80 | -12.09 | 1.23E-33 | Agree |
| Educational Attainment | Some College | -26.08 | 8.27 | -3.15 | 1.61E-03 | -10.26 | 0.77 | -13.35 | 1.30E-40 | Agree |
| Educational Attainment | College Degree | -49.07 | 8.17 | -6.01 | 1.91E-09 | -11.71 | 0.87 | -13.51 | 1.46E-41 | Agree |
| First-Degree Family History of Alzheimer's Disease | TRUE | -0.07 | 2.06 | -0.04 | 0.97 | 2.43 | 0.57 | 4.26 | 2.04E-05 | UK Biobank Finding |
| Age x Biological Sex | Men's vs. Women's per year change in Age slope | -0.62 | 0.26 | -2.38 | 2.00E-02 | -0.55 | 0.05 | -10.99 | 4.18E-28 | Agree |
| Biological Sex by Educational Attainment | Men with High School Diploma vs. Women with No High School Diploma | 21.63 | 19.62 | 1.10 | 0.27 | -11.24 | 1.61 | -6.96 | 3.30E-12 | UK Biobank Finding |
| Biological Sex by Educational Attainment | Men with Some College vs. Women with No High School Diploma | 16.25 | 18.28 | 0.89 | 0.37 | -10.54 | 1.54 | -6.83 | 8.71E-12 | UK Biobank Finding |
| Biological Sex by Educational Attainment | Men with a College Degree vs. Women with No High School Diploma | 14.79 | 17.94 | 0.82 | 0.41 | -12.8 | 1.74 | -7.37 | 1.68E-13 | UK Biobank Finding |

**Figure 4.**
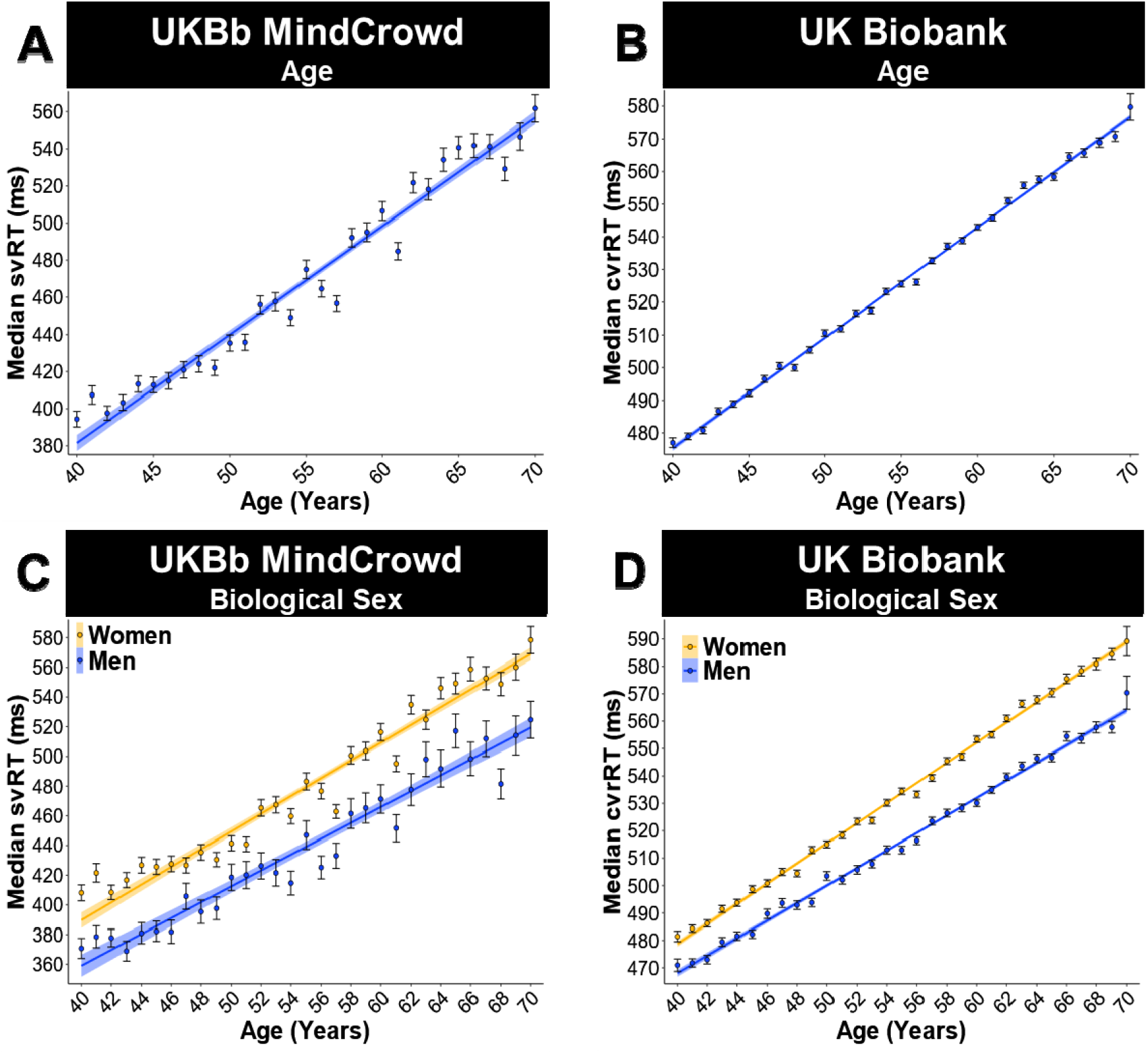
***<u>A-B</u>***<u>Age was linearly associated with visual reaction time (RT).UK Biobank analysis (ages 40-70).</u> (A) UKBb MindCrowd linear model fits (line fill ± 95% *CI*, error bars ± *SEM*) of median simple visual RT(svRT) across Age. (B) UK Biobank linear model fits (line fill ± 95% *CI*, error bars ± *SEM*) of mediancomplex, visual, recognition RT (cvrRT) across Age. UKBb MindCrowd svRT (β_*Age*_ = 5.75, *p*_*Age*_ = 2.00E-16, *n*= 39,795) and UK Biobank cvrRT (β_*Age*_ = 3.40, *p*_*Age*_ = 2.00E-16, *n* = 158,245) showed similar slowing from younger to older ages, but overall higher RT. The average 50 msec difference between UKBb MindCrowd svRT (*M* = 478.66 msec) and UK Biobank (*M* = 528.74 msec) is likely due to the choice component (i.e., do cards match or not > press button) of the UK Biobank’s cvrRT task compared to UKBb MindCrowd’s s imulus-response (i.e., the pink sphere appears > press button) svRT. ***<u>C-D</u>***<u>Visual reaction time (RT) was faster in men. UK Biobank analysis (ages 40-70).</u> (C) UKBb MindCrowd linear model fits (line fill ± 95% *CI*, error bars ± *SEM*) of median simple visual RT (svRT) across Age with lines split by Biological Sex. (D) UK Biobank linear model fits (line fill ± 95% *CI*, error bars ± *SEM*) of median complex, visual, recognition RT (cvrRT) across Age with lines split by Biological Sex. Both UKBb MindCrowd svRT (β_*Sex*_ = -40.00, *p*_*Sex*_ = 8.03E-71, 20.46%, Women [*M* = 489.75 msec, *n =* 29,640], Men [*M* = 446.28 msec, *n* = 10,155]) and UK Biobank cvrRT (β_*Sex*_ = -18.28, *p*_*Sex*_ = 2.00E-16, 5.16%, Women [*M* = 534.98 msec, *n =* 89,331], Men [520.66 msec, *n =* 68,91men consistently outperforming women from 40-70 years of age.) found

**Figure 5.**
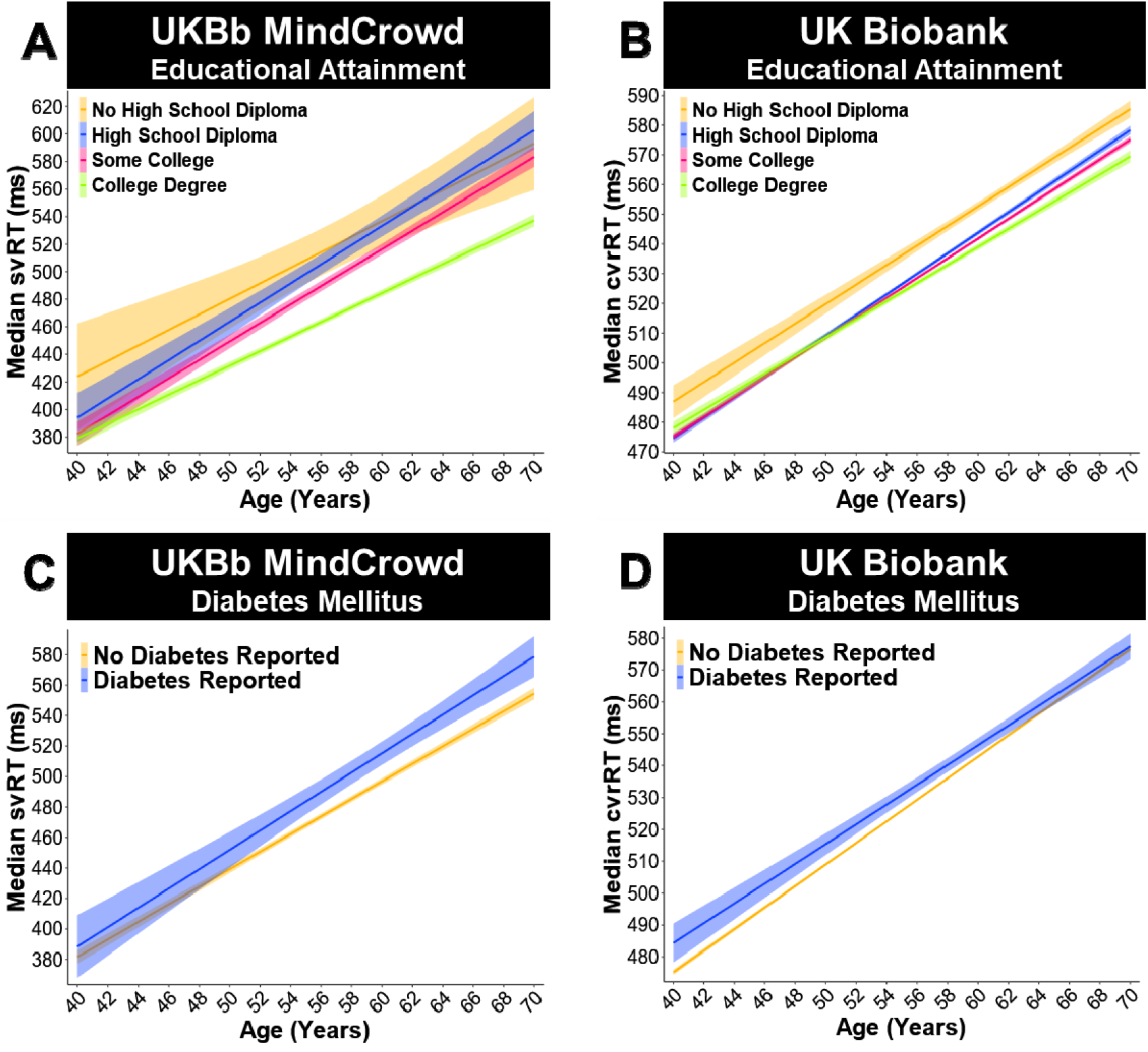
***<u>A-B</u>***<u>More education was related to faster visual reaction time (RT).</u> UK Biobank analysis (ages 40-70). (A) UKBb MindCrowd linear model fits (line fill ± 95% *CI*) of median simple visual RT (svRT) across Age with lines split by Educational Attainment. Participants who had a “High School Diploma” (β_*HSDiploma*_ *=* -9.68, *p*_*HSDiploma*_ = 2.70E-01, 1.87%, *n* = 3,176), “Some College” (β_*College*_ = -26.08, *p*_*College*_ = 1.61E-03, 5.06%, *n=* 11,139), or a “College Degree” (β_*CDegree*_ = -49.07, *p*_*CDegree*_ = 1.91E-09, 9.51%, *n =* 24,875) were faster than those with “No High School Diploma” (*n =* 605). (B) UK Biobank linear model fits (line fill ± 95% *CI*) of median complex, visual, recognition RT (cvrRT) across Age with lines split by Educational Attainment. Like the UKBb MindCrowd cohort, participants who had a “High School Diploma” (β_*HSDiploma*_ = -9.68, *p*_*HSDiploma*_ =1.23E-33, 1.74%, *n =* 46,247), “Some College” (β_*College*_ = -10.26, *p*_*College*_ = 1.30E-40, 1.85%, *n =* 77,2 0), or a“College Degree” (β_*CDegree*_ = -11.71, *p*_*CDegree*_ = 1.46E-41, 2.11%, *n =* 23,750) were faster than those with “No High School Diploma” (*n =* 10,978). ***<u>C-D</u>***<u>Diabetes mellitus slowed visual reaction time (RT). UK Biobank analysis (ages 40-70).</u> (C) UKBb MindCrowd linear model fits (line fill ± 95% *CI*) of median simple visual RT (svRT) across Age with lines split by diabetes mellitus. (D) UK Biobank linear model fits (line fill ± 95% *CI*) of median complex, visual, recognition RT (cvrRT) across Age with lines split by diabetes mellitus. Participant visual RT performance for UKBb MindCrowd svRT (β_*Diabetes*_ = 11.48, *p*_*Diabetes*_ = 3.31E-03, Diabetes Reported *n=* 2,807, No Diabetes Reported *n =* 36,988) and UK Biobank cvrRT (β_*Diabetes*_ = 5.48, *p*_*Diabetes*_ = 4.80E-07, Diabetes Reported *n =* 4,969, No Diabetes Reported *n =* 153,276), was slowed with Diabetes Mellitus.

### Age and biological sex

The UKBb MindCrowd cohort revealed Age as a significant predictor of svRT (*p*_*Age*_ = 2.00E-16). The parallel analysis (see Statistical Methods) of Age in the UK Biobank cohort was also significant (*p*_*Age*_ = 2.00E-16) for complex, visual, recognition reaction time (cvrRT). For the association of Age and RT, UKBb MindCrowd and the UK Biobank showed longer RTs or worse RT performance from younger to older ages, with nearly 6 and 3 msec slower RT per year difference of age, respectively (Figure 4A-B). For UKBb MindCrowd, Biological Sex was a significant predictor of RT (β_*Sex*_=-40.00, *p*_*Sex*_=8.03E-71), which was also the case in the UK Biobank (β_*Sex*_=-18.28, *p*_*Sex*_=2.00E-16). Being a man in both cohorts was associated with shorter RTs compared to being a woman (Figure 4C-D). Here the effect of Biological Sex on RT between UKBb MindCrowd was 40 msec (20.46%), and the UK Biobank was 18 msec (5.16%).

### Education and handedness

Akin to the association found in the MindCrowd analysis, the UKBb MindCrowd and UK Biobank comparison found that Educational Attainment was a significant RT predictor. Indeed, for UKBb MindCrowd a “High School Diploma” (β_*HSDiploma*_ *=* -9.68, *p*_*HSDiploma*_ = 2.70E-01, 1.87%, “Some College” (β_*College*_ = -26.08, *p*_*College*_ = 1.61E-03, 5.06%) and a “College Degree” (β_*CDegree*_ = -49.07, *p*_*CDegree*_ = 1.91E-09, 9.51%), and in the UK Biobank a “High School Diploma” (β_*HSDiploma*_ = -9.68, *p*_*HSDiploma*_ = 1.23E-33, 1.74%), “Some College” (β_*College*_ = -10.26, *p*_*College*_ = 1.30E-40, 1.85%), or a “College Degree” (β_*CDegree*_ = -11.71, *p*_*CDegree*_ = 1.46E-41, 2.11%) were all significantly different from “No High School Diploma” (Figure 5A-B). Here, both UKBb MindCrowd and UK Biobank large cohorts reported shorter RT was associated with more education. Lastly, unlike the MindCrowd analyses, both the UKBb MindCrowd *(p*_*Handedness*_ = .40) and the UK Biobank (*p*_*Handedness*_ = 0.36) cohorts between the ages of 40-70 did not find Handedness to be a significant predictor of RT performance (Supplementary Figure 6).

**Figure 6.**
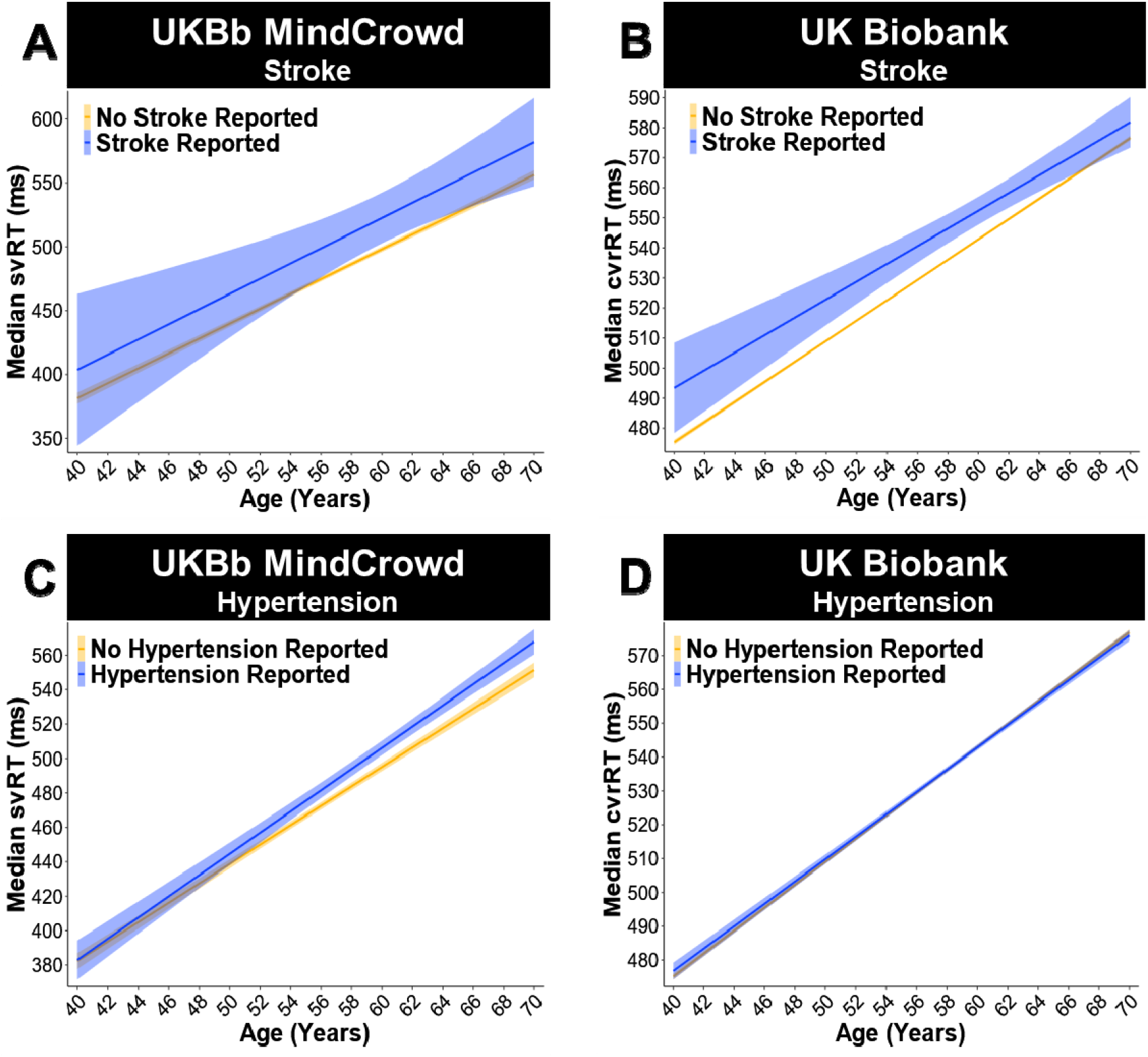
***<u>A-B</u>***<u>Visual reaction time (RT) was impaired if a Reported Stroke was reported. UK Biobank analysis (ages 40-70).</u> (A) UKBb MindCrowd linear model fits (line fill ± 95% *CI*) of median simple visual RT (svRT) across Age with lines split by Reported Stroke. (B) UK Biobank linear model fits (line fill ± 95% *CI*) of mediancomplex, visual, recognition RT (cvrRT) across Age with lines split by Reported Stroke. In both th UKBbMindCrowd svRT (β_*Stroke*_ = 18.47, *p*_*Stroke*_ = 4.00E-02, Reported Stroke Reported *n =* 2,807, No Reported Stroke Reported *n =* 39,318) and UK Biobank cvrRT (β_*Stroke*_ = 10.61, *p*_*Stroke*_ = 6.15E-07, Reported Stroke Reported *n =* 1,237, No Reported Stroke Reported *n =* 157,008) analysis, experiencing a Reported Stroke was associated with slower visual RT. ***<u>C-D</u>***<u>Reported Hypertension slowed both types of visual reaction time (RT). UK Biobank analysis (ages 40-70).</u> (C) UKBb MindCrowd linear model fits (line fill ± 95% *CI*) of median simple visual RT (svRT) across Age with lines split by Reported Hypertension. (D) UK Biobank linear model fits (line fill ± 95% *CI*) of median complex, visual, recognition RT (cvrRT) across Age with lines split by Reported Hypertension. Unlike the MindCrowd analysis, hypertension was related to slower svRT in UKBb MindCrowd (β_*Hypertension*_ = 7.99, *p*_*Hypertension*_ = 3.16E-03, Hypertension Reported *n =* 9,676, No Hypertension Reported *n =* 30,119) andcvrRT in the UK Biobank (β_*Hypertension*_ = 1.14, *p*_*Hypertension*_ = 0.02, Hypertension Reported *n =* 32, Hypertension Reported *n =* 125,652).93, No

### Health, medical, and lifestyle factors

In terms of health factors associated with RT, in the UKBb MindCrowd cohort, Diabetes (β_Diabetes_=11.48, *p*_*Diabetes*_=3.31E-03, 5.87%), Stroke (β_*Stroke*_=18.47, *p*_*Stroke*_=4.00E-02, 9.45%), (β_*Hypertension*_=7.99, *p*_*Hypertension*_=3.16E-03, 3.58%), and Dizziness (β_*Dizzy*_=12.13, *p*_*Dizzy*_=2.52E-03, 6.19%) were all significantly associated with longer svRTs. These associations were recapitulated by the UK Biobank. To that end, Diabetes Mellitus (β_*Diabetes*_ = 5.48, *p*_*Diabetes*_ = 4.80E-07, 1.55%, Figure 5C-D), Reported Stroke (β_*Stroke*_ = 10.61, *p*_*Stroke*_ = 6.15E-07, 2.99%, Figure 6A-B), Reported Hypertension (β_*Hypertension*_ = 1.14, *p*_*Hypertension*_ = 0.02, 0.31%, Figure 6C-D), and Reported Dizziness (β_*Dizzy*_ = 3.21, *p*_*Dizzy*_ = 3.71E-14, 0.91%, Figure 7A-B) were all significantly related to longer cvrRTs; however, the association between Reported Hypertension and cvrRT was small (Figure 6D). Further, in agreement with the MindCrowd analysis, Smoking Status was significantly related to svRT (β_*Smoke*_=10.18, *p*_*Smoke*_=0.01, 5.21%) in the UKBb MindCrowd cohort; however, FHAD (β_*FHAD*_=-0.07, *p*_*FHAD*_=0.97) was not. This pattern of associations were reversed in the UK Biobank; that is, Smoking Status was not a significant predictor of cvrRT (β_*Smoke*_=0.47, *p*_*Smoke*_=0.22, Figure 7C-D), but FHAD was (β_*FHAD*_=2.36, *p*_*FHAD*_=3.35E-05, 0.69%, Figure 8A-B).

**Figure 7.**
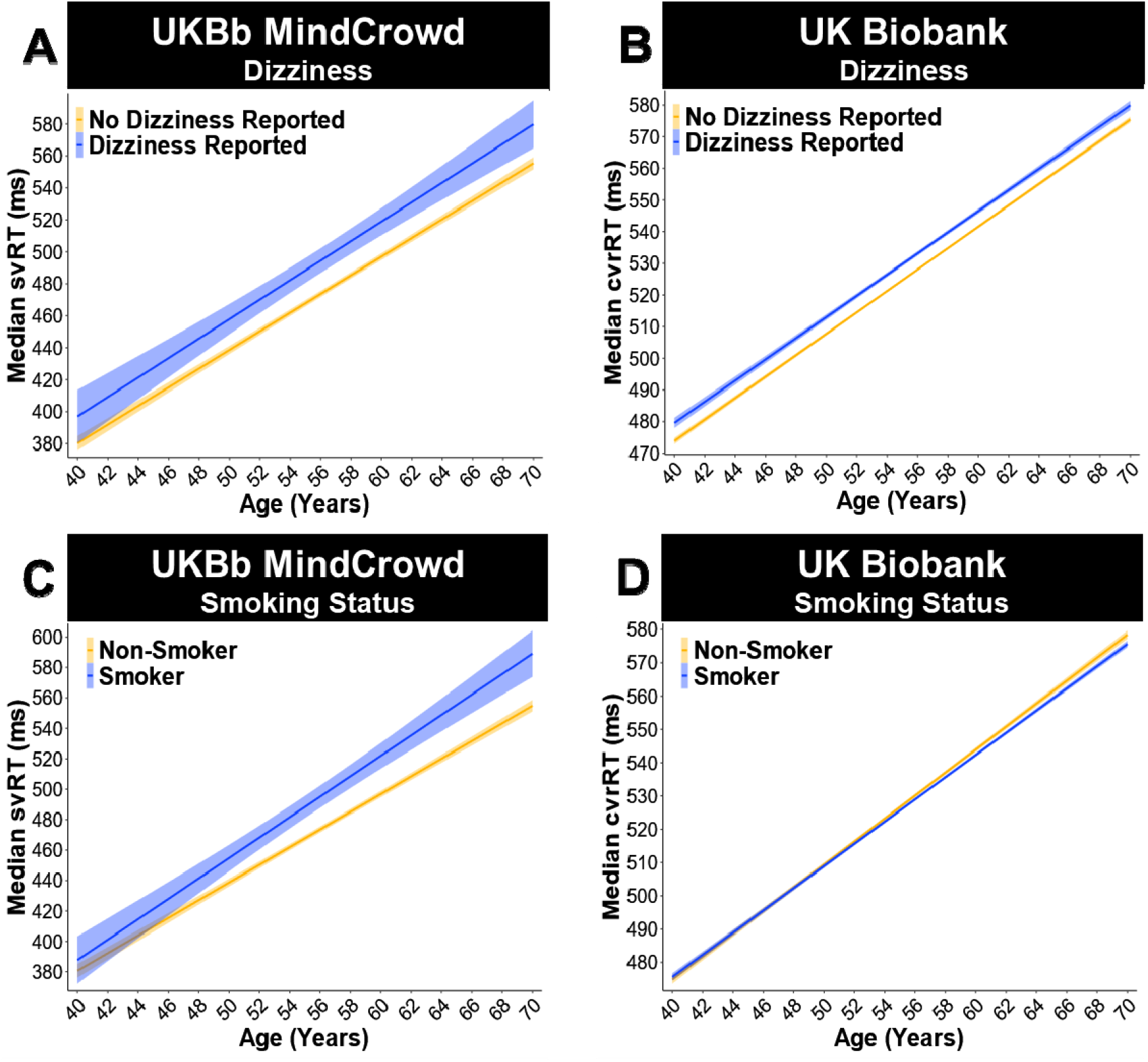
***<u>A-B</u>***<u>Reported Dizziness impaired visual reaction time (RT). UK Biobank analysis (ages 40-70).</u> (A) UKBb MindCrowd linear model fits (line fill ± 95% *CI*) of median simple visual RT (svRT) across Age with lines split by Reported Dizziness. (B) UK Biobank linear model fits (line fill ± 95% *CI*) of median complex, visual, recognition RT (cvrRT) across Age with lines split by Reported Dizziness. UKBb MindCrowd svRT (β_*Dizzy*_ = 12.13, *p*_*Dizzy*_ = 2.52E-03, Reported Dizziness Reported *n =* 2,543, No Reported Dizziness Reported *n =* 37,252) and UK Biobank cvrRT (β_*Dizzy*_ = 3.21, *p*_*Dizzy*_ = 3.71E-14, Reported Dizziness Reported *n =* 42,210, No Reported Dizziness Reported *n =* 116,035) were slowed if participants reported Reported Dizziness. C-D <u>Smoking Status is related to impaired MindCrowd simple visual reaction time (svRT), not UK Biobank complex, visual recognition reaction time (cvrRT).</u> UK Biobank analysis (ages 40-70). (C) UKBb MindCrowd linear model fits (line fill ± 95% *CI*) of median svRT across Age with lines split by Smoking Status. Compared to non-smokers, smokers demonstrated slower svRT (β_*Smoke*_ = 10.18, *p*_*Smoke*_ = 0.01, Smoker *n =* 2,783, Non-Smoker *n =* 37,012). (D) UK Biobank linear model fits (line fill ± 95% *CI*) of median cvrRT across Age with lines split by Smoking Status. An association between Smoking Status and cvrRT was not found in the UK Biobank (β_*Smoke*_ = 0.47, *p*_*Smoke*_ = 0.22, Smoker *n =* 91,312, Non-Smoker *n =* 66,923).

### Two-way interactions

The “*glmulti*” ^48^ R package defined two interactions in the UKBb MindCrowd and UK Biobank analysis. In UKBb MindCrowd we found a significant Age x Biological Sex interaction (*p*_*Age*Sex*_ = 2.00E-02, 0.33%). We found a comparable significant Age x Biological Sex interaction for cvrRT (*p*_*Age*Sex*_ = 4.18E-28, 0.16%) in the UK Biobank. Across both MindCrowd and the UK Biobank, these interactions indicated that RT was lengthened in men from younger to older ages compared to women, was over 0.5 msec shorter per one year difference in age (Figure 4C-D). In addition, the UK Biobank analysis revealed a significant Biological Sex x Educational Attainment interaction not found in UKBb MindCrowd. Here, men with a “High School Diploma” (*p*_*Sex*HSDiploma*_ = 3.30E-12, 3.35%), “Some College” (*p*_*sex*College*_ = 8.71E-12, 3.14%) or a “College Degree” (*p*_*sex*CDegre*_ = 1.68E-13, 3.18%) were significantly associated with faster cvrRT performance when compared to women having “No High School Diploma” (Figure 8C). Follow-up analyses of the simple effects via estimated marginal means (EMM, see Statistical Methods) revealed that men who did not graduate high school (*EMM* = 558.91 msec), compared to men with more education (*EMMs* = 542.99, 542.98, and 540.41 msec), had markedly faster cvrRTs, more in line with the women’s cvrRT performance (*EMMs =* 566.80, 562.11, 561.40, and 561.09 ms). The associated difference in cvrRT for men with “No High School Diploma” compared to men with a “High School Diploma” (β_*Men*_*=*15.92, *p*_*Men*_ = 2.00E-16, 1.75%) was more substantial than between women with “No High School Diploma” compared to women with a “High School Diploma” (β_*Women*_*=*4.68, *p*_*Women*_ = 3.12E-04, 1.69%, see Supplementary Figure 7 for a graph of the EMM).

## Discussion

### Slowing of Reaction Time (RT) from Younger to Older Ages: Factors and Modifiers

Our study’s results illuminate a portion of the intricate relationship between age and RT performance by identifying demographic, health, medical, and lifestyle factors associated with either attenuation or exacerbation of RT lengthening from younger to older ages (see figure 9 for an illustrative summary of the results). A large body of work on RT, leading back to Sir Francis Gaulton in 1890^49^, has consistently demonstrated an age-associated shift in RT^50^. It is not surprising that MindCrowd, UKBb MindCrowd, and UK Biobank models revealed slowing of simple visual (svRT, MindCrowd) and complex visual recognition RT (cvrRT, UK Biobank) from younger to older ages. Likewise, the MindCrowd model found that the relationship between svRT and age was modestly curvilinear (Figure 1A). While this curvilinear relation between RT and age has been noted previously^51^, both cohorts’ large sample size combined with our application of an algorithm-based model definition^48^ revealed a notable addition to this picture. Specifically, in the MindCrowd analysis, we observed an interaction between age and education (Figure 2A) and smoking (Figure 3A). Here, less education and smoking were related to the additional slowing of simple visual RT (svRT) on top of the svRT slowing associated with transitioning from younger to older ages. Lastly, the age by reported stroke interaction modeled in MindCrowd was associated with longer RTs (Figure 3E). This study’s large sample size and broad age and surveyed data range places it as one of the most substantial cross-sectional RT evaluations across the aging spectrum. Our findings suggest that smoking and stroke (i.e., cardiovascular health) and amount of education (i.e., cognitive demand or reserve) are factors, modifiable across aging, that influence age-associated RT slowing.

### A First-Degree Family History of Alzheimer’s Disease (FHAD), Cognition, and RT: Intertwined systems

In the UK Biobank cohort^27^, we found an association between having an FHAD and slower (2.43 msec) cvrRT (Figure 8B). This effect of FHAD on RT or more so underlying process speed is in line with our episodic verbal memory task (i.e., paired-associate learning [PAL]) finding ^34^, where we found FHAD was linked to lower PAL performance. Furthermore, a prior functional magnetic resonance imaging (fMRI) study examining medial-temporal lobe activation using a cvrRT task found a ∼100 msec RT lengthening in 68 (mean age of 54) FHAD participants^52^. These data suggest that genetic and environmental factors relating to AD risk are present in individuals with an FHAD. Indeed, the first-degree relatives of identified FHAD participants consisted of familial early-onset AD or late-onset AD, which also has a high heritability of 79%, suggesting that they are a higher risk category for developing AD. Thus, such shared AD and FHAD factors may relate to sensorimotor function and processing speed (i.e., RT) analogous to alterations in cognition and memory (i.e., PAL).

We found a correlation between svRT and PAL performance (Figure 1B). This finding was in line with many prior studies, the dependence of episodic memory on processing speed, a dependence that grows with age and incident of age-related disease (e.g., AD)^5-8,53^. The association of svRT with PAL may highlight distributed systems and networks that underlie RT performance. For example, svRT performance could depend on the functioning of many cognitive areas that PAL requires and vice versa. Another possibility is that properly functioning memory, and cognitive networks correspond to better RT performance. Evidence for this is suggested by the fact that higher intelligence is related to faster RT^54-56^. However, diverse factors (e.g., exercise, time of day, meal proximity, and individual assessing RT) affect RT performance^53,56,57^. Thus, making an accurate estimation of RT’s effect on intelligence and vice versa changeling. Together with our prior results, these findings suggest that RT performance could be used as a metric to assess potential AD risk. However, further research, including longitudinal studies, replication, and corroboration of RT’s link to age-related cognitive decline and disease, are necessary to support this notion.

The difference in the effects of FHAD between UK Biobank and MindCrowd (for both MindCrowd and UKBb MindCrowd analyses, Supplementary Figure 3A and Figure 8A) could be due to the vast difference in the fraction of FHAD participants in the UK Biobank (FHAD = 13% of total) compared to UKBb MindCrowd (FHAD = 35% of total). While noting that we target those with FHAD for recruitment into MindCrowd, this substantial disparity could also be due to the accuracy of the UK Biobank’s FHAD. The UK Biobank was calculated from three separate questions (i.e., mother, father, and siblings AD diagnosis). Adding to this, the United Kingdom uses a massive, detailed, nation-wide electronic health record system facilitating respondent health and medical survey accuracy. Compare this to our single question in MindCrowd, asking participants of all ages to remember if a relative was diagnosed with AD. Another possibility is the RT paradigm used; that is, MindCrowd’s test of svRT compared to the UK Biobank’s use of cvrRT. The fact that complex reaction time, requiring recognition and the choice to “respond or not respond,” rather than just stimulus-response, may underly this difference. UKBb MindCrowd svRT performance was consistently faster across age than UK Biobank cvrRT performance; an observation noted in a prior study also measured both simple and complex RT^58^. These findings are consistent with the idea that complex RT requires more processing time^12,59^, and prior work found the age-associated slowing of RT was higher for choice RT than simple RT ^60^. Lastly, the differences in FHAD associations across MindCrowd and the UK Biobank are perhaps due to the UK Biobank had over 2X the number of participants compared to MindCrowd (i.e., 158K vs. 76K). The larger sample is expected to produce better model definitions and increased statistical power. Increased statistical power may also have enhanced accuracy, validity, and reproducibility.

### Biological Sex and Educational Attainment

Numerous RT studies have found sex differences in RT performance, which does not appear to be reduced by practice^16,20,58^. Consistent with others^58,60^, men exhibited shorter RTs in each model across cohorts (Figures 1D and 4C-D). In addition, the analysis of the UKBb MindCrowd and UK Biobank implicated biological sex affecting RT slowing from younger to older ages. The age interaction with biological sex suggests that being a woman from younger to older ages is associated with longer RT compared to being a man. These results largely replicate a previous sizable study (i.e., 7,000 participants) evaluating RT^58^. Similar to our own, this study found that 1) men consistently outperformed women on all RT measures across age, 2) differences in RT performance from younger to older ages were nonlinear, 3) including a third-degree polynomial for age provided the best model fit, and 3) compared to men, women displayed longer RTs consistently from younger to older ages^58^. Collected with our prior study of PAL performance ^34^, these associations replicate prior work and suggest that biological sex affects RT and age-associated shift in RT.

Educational attainment was associated with svRT in both MindCrowd cohorts and cvrRT in the UK Biobank. Overall, having more education (i.e., reporting higher milestones) was related to shorter svRT and cvrRT (Figures 2A and 5A-B). However, it is unclear if individuals with higher processing speed naturally seek more education and what other factors confound this relationship. Further work utilizing both cohorts is necessary to shed light on the effects and modifiers of FHAD and cross-cohort discrepancies. The model of the UK Biobank revealed an interaction between biological sex and education on RT performance. The breakdown revealed that men had similar RT performance if they attained a high school diploma and above. However, men who did not graduate high school showed markedly slower RTs, which brought them in line with women’s RT performance. However, the associated lengthening of RT for less-educated men was vastly more than that found in less-educated women (reported “No High School Diploma” made up 3.83% of women and 3.11% of men; see Supplementary Figure 7).

### Handedness

In MindCrowd, handedness, specifically being left-handed, was associated with shorter svRTs. Prior studies have reported similar associations, where left-handedness was correlated with shorter svRT^19,59,61^. Hemispheric asymmetries in spatial processing are thought to underly faster svRT for the left hand^60,62^. Handedness was not associated with svRT in UKBb MindCrowd or cvrRT in the UK Biobank. One explanation for the divergent findings is that MindCrowd includes younger participants (i.e., 18–40-year-olds). Indeed, in MindCrowd, the association appears to diminish from younger to older ages. Specifically, in Figure 2B, the separation of the regression lines between left- and right-handed participants shrinks and eventually crosses around the 4^th^ decade of life. Figure 2C shows that the left- and right-handed regression lines separate in 20 to 40-years-olds, while Figure 2D shows that these regression lines are not separate in 40 to 60-year-olds. While purely speculative, differences in social conventions may have played a role. For example, some older participants were forced to be right-handed, whereas younger participants were not. In doing so, upping the amount of unexplained variance in older, but not younger, participants across MindCrowd and the UK Biobank.

### Health, Medical, and Lifestyle Factors

The MindCrowd analysis incorporated all 13 available health, medical, and lifestyle-related factors, of which six were present and incorporated into the shared UKBb MindCrowd/UK Biobank model (Tables 2 and 4). Before the launch of MindCrowd, these factors were carefully selected based upon their known relation to 1) age-associated alterations, 2) RT performance, and 3) PAL Performance. Of the 13 health, medical, and lifestyle factors evaluated in the MindCrowd analysis, we found associations between svRT and the number of daily medications, reported dizziness, smoking status, reported stroke, and diabetes mellitus. Each health and medical factor were associated with longer svRTs (Figure 3). We should note that the number of daily medications is a serving as a proxy for overall health. That is, the worse one’s health, the worse one’s performance, the increased number of medications treating the underlying health conditions. The UKBb MindCrowd (svRT) and the UK Biobank (cvrRT) analyses found *similar* associations between reported dizziness, reported stroke, diabetes mellitus, indicating hypertension. Although each association differed in magnitude between the two older cohorts, each was related to slower RT. The UKBb MindCrowd to the UK Biobank found a different association for FHAD (UKBb MindCrowd = no association; UK Biobank = 2.43 msec slower), smoking status (UKBb MindCrowd = 10 msec slower; UK Biobank = no association). Interestingly, despite some differences, only a few coefficient signs differed between the UKBb MindCrowd and UK Biobank; indeed, most estimations were well within an order of magnitude between the two cohorts (e.g., age, educational attainment, and age by biological sex interaction, Table 5).

Many factors are likely to account for the different associations between smoking and FHAD between UKBb MindCrowd and the UK Biobank (Figures 7C-D and 8A-B). Some of these include differences in demographics, genetic heterogeneity, and age^26^. However, candidates include the fractions of participants reporting each factor (e.g., for diabetes mellitus: MindCrowd = 1%, UKBb MindCrowd = 7%, and UK Biobank = 3%). Another factor is that the UK Biobank’s participant number is twice the size of MindCrowd and four times the size of UKBb MindCrowd. Despite our study’s size, the observational and cross-sectional method means that we cannot rule out effects due to confounding variables.

Consequently, while numbers may be close, we do not assume that the UKBb MindCrowd is similar and can be compared to the UK Biobank. Furthermore, we observed that UKBb MindCrowd consistently reported larger estimates and standard errors than the UK Biobank. For example, the MindCrowd cohort’s estimation of the sex-difference was consistently slower (∼40 msec), even in the UKBb MindCrowd cohort, when looking at the UK Biobank (∼19 msec). This difference demonstrates why the study of neuropsychological traits and disease requires large sizes to provide accurate estimations driving better predictive validity.

### Opportunities and Limitations of Our Research

### Opportunities: Unique impact

We strongly advocate for large-scale efforts like ours, the UK Biobank^63^, and others^22^. Indeed, studies of this kind have characteristics that provide the unique impact necessary to move the fields of aging and age-related diseases forward. These include: 1) statistical control, as our MindCrowd analysis incorporated all 24 available factors, 11 of which were used in the UKBb MindCrowd and UK Biobank model. 2) The inclusion of each predictor controlled for its association on RT, which potentially removed variability (noise), thus enhancing statistical power. 3) The two models used for each of the analyses were selected with little human input by automated application of specific statistical criteria (see Inclusion of polynomials and automatic model selection in Statistical Methods). This likely decreases bias, the probability of overfitting, and multicollinearity. 4) For the present study, MindCrowd had over 76K and the UK Biobank over 158K participants. Large sample sizes in each cohort were expected to help reduce variance, enhance estimation, select better models, and in turn, enhance statistical power. Expanded statistical power may then enhance accuracy, validity, and reproducibility.

Lastly, a recent genome-wide association study examining associations between RT and single nucleotide polymorphisms (SNP) in the UK Biobank and CHARGE and COGENT consortia noted weak correlations between the reported cognitive-associated SNPs among US and UK cohorts^64^. Here, MindCrowd presents a future opportunity to resolve these weak associations and get a better picture of potential cohort effects. Taken together, these characteristics increase the likelihood of making accurate inferences regarding associations while boosting predictive validity. These are both necessary and vital attributes when searching for genetic associations and the structure underlying healthy brain aging.

### Limitations: Internet self-report and others

There are potential concerns that arise from web-based studies^65^. Indeed, limitations of this study include the cross-sectional design and the partial discrepancy in MindCrowd’s svRT test compared to the UK Biobank’s cvrRT test and info collected between the UK Biobank and MindCrowd (e.g., the omission of “prefer not to answer” choices for race and education questions). Acknowledging these drawbacks, we believe that the advantage of meaningfully higher participant numbers and enriched cohort diversity facilitated via online research remediates some disadvantages. For example, the range of error reported in recent internet-based studies of self-reported quantitative traits like height and weight was between 0.3% and 20% ^66-69^. Previously, we ran simulations on the association between FHAD and PAL by randomly shuffling the FHAD responses (e.g., Yes to No, and No to Yes), introducing increasing sequential amounts of “error.” We found that even with a subtle effect such as FHAD on PAL performance, due to our cohort size, 24% error would still have only made us commit a Type1 error 50% of the time^34^. In line with this notion, another publication demonstrated that online RT studies produce reproducible results^70^.

Further, we developed an extensive and automated data filtering pipeline (see Data Quality Control and Supplementary Figures 9-10) to address these concerns and enhance validity and accuracy. For example, we controlled for svRT delays generated using the touchscreen or mobile device by adding this information as a variable to the regression model (see Supplementary Figure 8). These data (i.e., real or filtered) were excluded before analysis (i.e., listwise deletion). Exclusion resulted in dropping 0.3 % and 6.1% of MindCrowd and UK Biobank participants, respectively. One of the 25 critical factors had over 5% missing data (see Supplementary Figure 11 and Supplementary Tables 4 and 5). Reported Dizziness in the UK Biobank had 64.36% missing data. Hence, interpretation of this factor’s association with cvrRT should only be considered for “hypothesis-generation^71^.” Evaluation of selection bias between retained and excluded participants revealed an overall lower probability of exclusion in MindCrowd and higher likelihood in the UK Biobank (see Supplementary Tables 6 and 7). Notable groups with a higher probability of exclusion included those in the highest age ranges and those reporting hypertension and dizziness. These higher probability groups were found in both study’s cohorts.

Lastly, it is essential to note that our internet-based svRT task was not designed to mirror conventional face-to-face RT testing paradigms directly. Indeed, we find higher RTs and steeper slopes across aging than studies assessing svRT via the gold standard, laboratory-based assessments (e.g., ^60,72^). However, these paradigm differences are not likely to alter our svRT test’s validity or reliability. One reason being our test is only interpreted within MindCrowd to identify associated factors and reveal individual differences. Despite test paradigm differences, we believe that large cross-sectional studies like MindCrowd, utilizing internet-based testing and remote biosample collection, are vital to moving the field of aging and age-related disease forward (see Opportunities: Unique impact above and ^63^).

## Conclusion

Understanding the modifiable and non-modifiable variables associated with RT and related cognitive function will begin to deconstruct the underlying architecture of elements accounting for the vast heterogeneity seen in individual trajectories of age-associated cognitive decline. Only then will it be possible to develop a healthy brain aging model that is both valid and reliable^73^. Such a model holds immense potential to attenuate age- and disease-related cognitive deficits, thus enhancing cognitive healthspan. Any extension of cognitive healthspan, better aligning it to the human lifespan, would be invaluable and increasingly vital when aggregated across the aging population. Mitigating age-or disease-related cognitive decline, allowing maintenance of independence by even only a few years, would have many benefits. For example, the U.S. could save billions of dollars in health care costs and lost productivity of caregivers while improving the quality of life for the aging population^63^. In this study, we revealed several potential factors related to aging and processing speed. Of those, smoking and education, as potentially modifiable factors throughout life, were associated with slower and faster RTs, respectively, from younger to older ages. With MindCrowd recruitment ever-increasing, our goal is to continue supplying and refining the knowledge necessary to optimize cognitive performance throughout life.

## Supporting information

Combined Supplemental Figures and Tables

## Data Availability

Final data will be filtered to exclude irrelevant data fields and be made available before peer-reviewed publication on Dryad (https://datadryad.org).

## Data Availability

All programs, software, and other materials described herein are publicly or commercially available. R code used for all analyses and figure generation was included as “Source Data” R Markdown notebook. The R code includes each model selected and the full list of parameters used. The data that support each analysis, figure, and table is freely available at Dryad (https://datadryad.org).

## Acknowledgments

A substantial porting of this research was conducted using the generous data and resources provided by the UK Biobank (https://www.ukbiobank.ac.uk/published-papers). We acknowledge the Mueller Family Charitable Trust and Mike Mueller their significant donations. The State of Arizona DHS also supported this work in support of the Arizona Alzheimer’s Consortium (PI; Eric Reiman), the Flinn Foundation (PI; Matt Huentelman), The McKnight Brain Research Foundation, NIH-NIA grant R01-AG049465 (PI; Carol Barnes), and NIH-NIA grants UH2 AG064706, U19 AG023122, U24 AG051129, and U24 AG051129-04S1 (PI; Nicholas J. Schork). We also recognize the support of many anonymous donors and institutional support of TGen, the City of Hope, and the University of Arizona.

## Author Contributions

J.S.T wrote and edited much of the manuscript, analyzed much of these data, and supervised several project management areas. J.S.T also contributed to data analysis, interpretation, and synthesis. M.D.DB oversaw much of the information technology needed for this study, analyzed a portion of these data, and supplied critical project support. M.A.N analyzed a part of these data, generated some figures, and provided critical statistical support. A.M.S. helped write and edit the manuscript as well as conduct critical portions of the statistical analyses. C.R.L. aided in writing and editing the manuscript, data analysis, and general project oversight. W.M.J. aided in writing and editing the manuscript, data analysis, and general project oversight. A.K.H wrote and helped write portions of the manuscript and serve as a primary editor. Further, A.K.H. gave critical early data analysis and project support as well as data interpretation and synthesis. T.R. edited the manuscript and provided critical clinical insight and project management support. B.E.L. edited the manuscript and gave vital clinical insight. S.H. edited the manuscript and supported data analysis. Y.B. edited the manuscript and provided critical clinical insight and project management support. R.D.B. was instrumental in planning, designing, and executing this research. N.J.S. collaborated to provide access to the UK Biobank and helped with data analysis as well as project insight generation. M.H. supplied critical early data analysis and project support, as well as data interpretation and synthesis. C.A.B. helped write and edit the manuscript. In addition, C.A.B. aided several aspects of study design and helped with overall project planning and data interpretation and synthesis. E.G., aided in writing and editing the manuscript and study design and data interpretation and synthesis. L.R. aided in writing and editing the manuscript and study planning, design, and data interpretation and synthesis. M.J.H. was the principal investigator of this project. M.J.H oversaw most study planning, design, data collection and analysis, and data interpretation and synthesis. MJH played a substantial role in writing and editing the manuscript.

