## Supplementary material for "Two Separate, Large Cohorts Reveal Potential Modifiers of Age-Associated Variation in Visual Reaction Time Performance": Combined Supplemental Figures and Tables

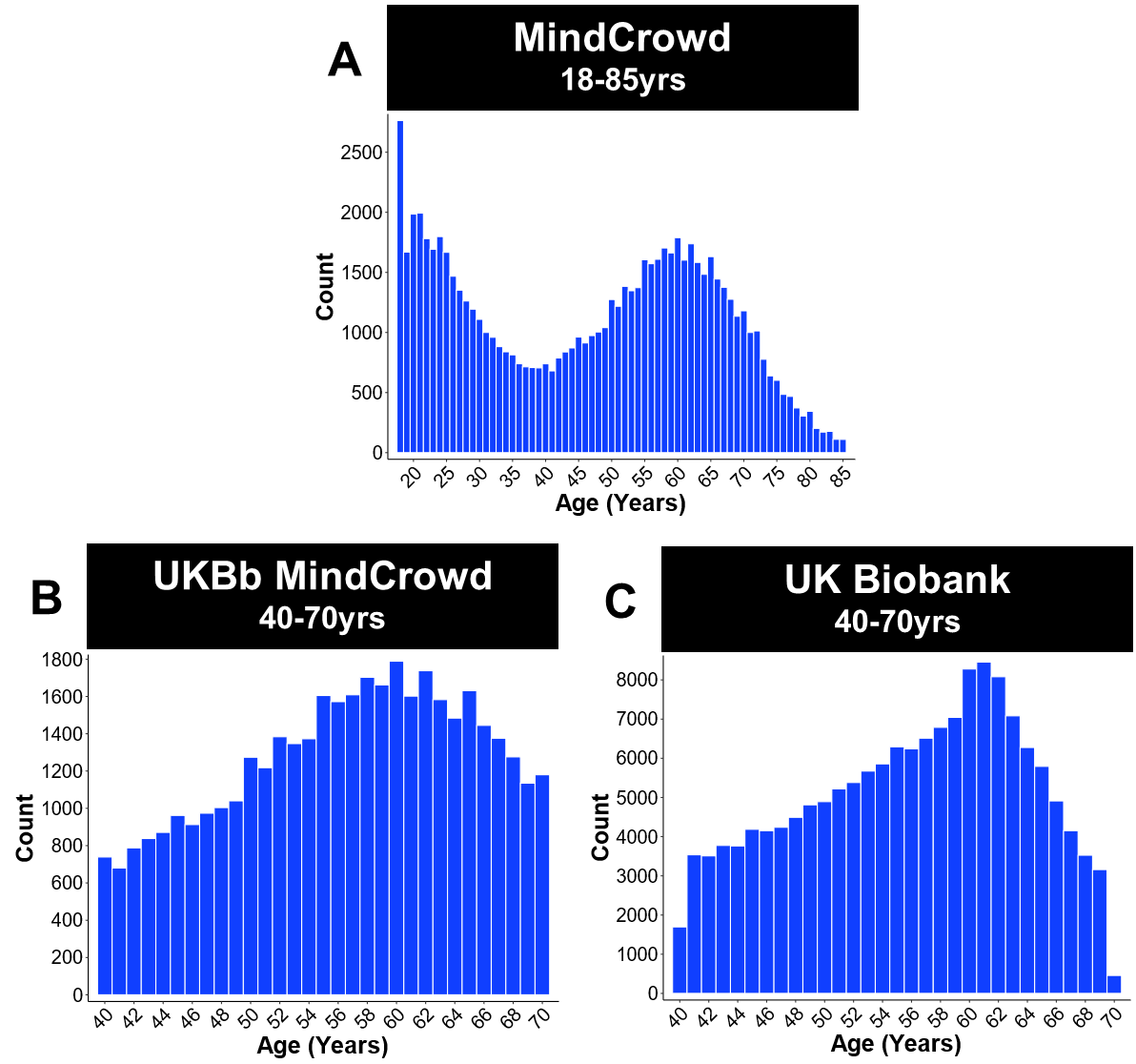
**Supplementary Figure 1.** Distribution of ages across MindCrowd and the UK Biobank. Histogram of age across a) MindCrowd (18-85 years, *n =* 73,464), b) UKBb MindCrowd (40-70 years, *n =* 38,891), and c) UK Biobank (40-70 years, *n =* 157,933) cohorts. Each bin is one year. These plots display the bimodal distribution of ages in the MindCrowd cohort and the similar and approximately normal distribution of the UKBb MindCrowd and UK Biobank cohorts.

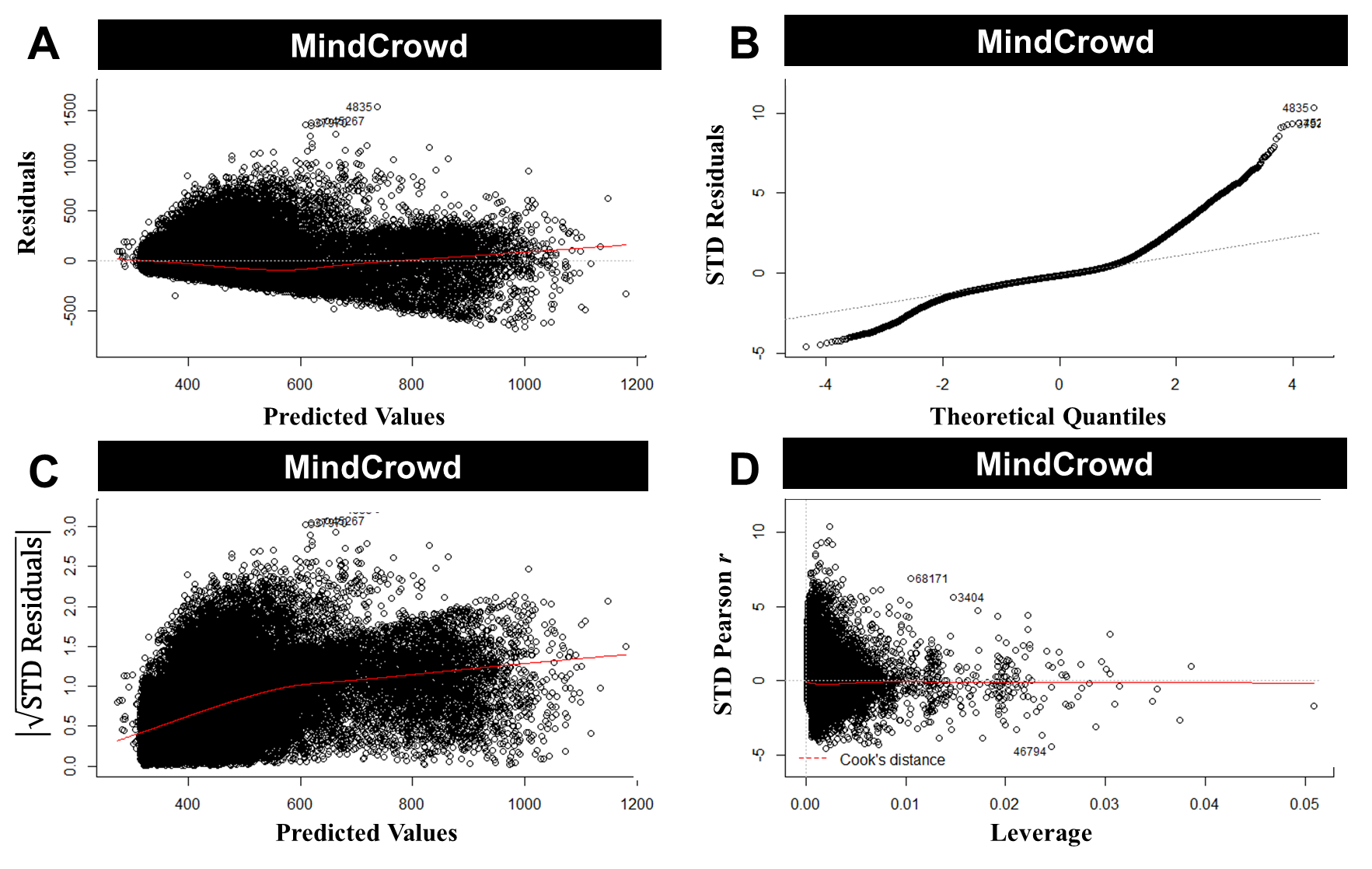
 **Supplementary Figure 2.** Regression diagnostic plots of the general linear model (LM) for the MindCrowd analyses (*n =* 75,666). (A) A plot of the residuals versus fitted values. This plot suggests that the assumptions of linearity, equality of variances, and no outliers are met. (B) Q-Q plot displaying the quantiles of the data versus quantiles of a normal distribution. This plot suggests a violation of normality in residuals and thus the error terms; however, this violation should not cause major problems because of the large sample size. (C) A plot of the absolute value of the standardized (STD) residuals’ square root versus the fitted values. Like plot A, this plot suggests variability in residuals does not change much over the range of the dependent variable. (D) A plot of the residuals versus leverage. This plot shows that there are no influential cases, as all cases fall within Cook’s distance.

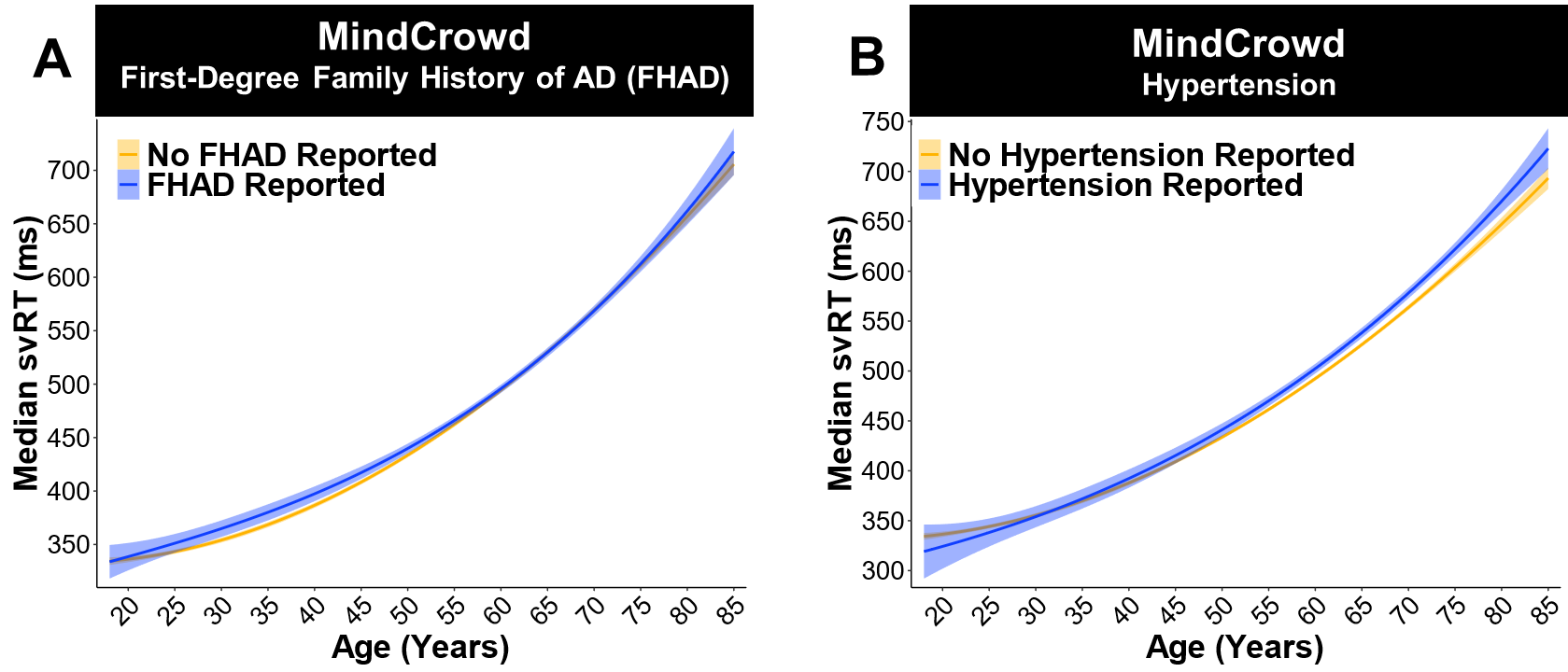

**Supplementary Figure 3.** MindCrowd: No relationship between a first-degree family history of Alzheimer’s disease (FHAD) or Reported Hypertension and simple visual reaction time (svRT). (A) MindCrowd analysis (ages 18-85): Linear model fits (line fill ± 95% *CI*) of the median svRT across Age with lines split by FHAD. There was no relationship between FHAD and svRT (*β_FHAD_* = -0.40, *p_FHAD_* = 0.75, FHAD Reported *n =* 17,847, No FHAD Reported *n =* 57,819). (B) Linear model fits (line fill ± 95% *CI*) of the median svRT across Age with lines split by Reported Hypertension. Hypertension was not related to svRT (*β_Hyper_* = 1.08, *p_Hyper_* = 0.52, Hypertension Reported *n =* 13,915, No Hypertension Reported *n =* 61,751).

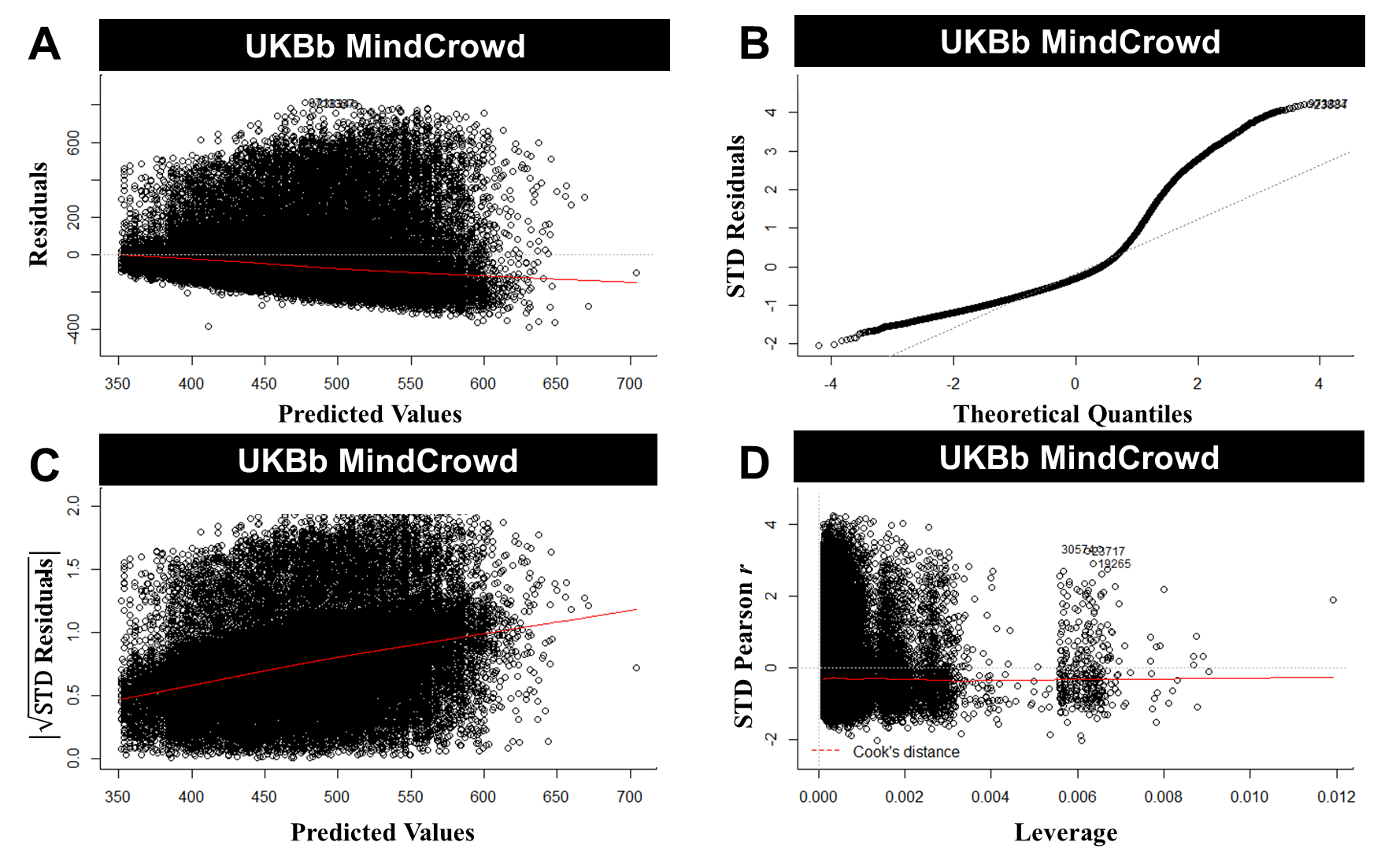
 **Supplementary Figure 4.** Regression diagnostic plots of the general linear model (LM) for the UKBb MindCrowd cohort (*n =* 39,795). (A) A plot of the residuals versus fitted values. This plot suggests that the assumptions of linearity, equality of variances, and no outliers are met. (B) Q-Q plot displaying the quantiles of the data versus quantiles of a normal distribution. This plot suggests a violation of normality in residuals and thus the error terms; however, this violation should not cause major problems because of the large sample size. (C) A plot of the absolute value of the standardized (STD) residuals’ square root versus the fitted values. Like plot A, this plot suggests variability in residuals does not change much over the range of the dependent variable. (D) A plot of the residuals versus leverage. This plot indicates that there are no influential cases, as all cases fall within Cook’s distance.

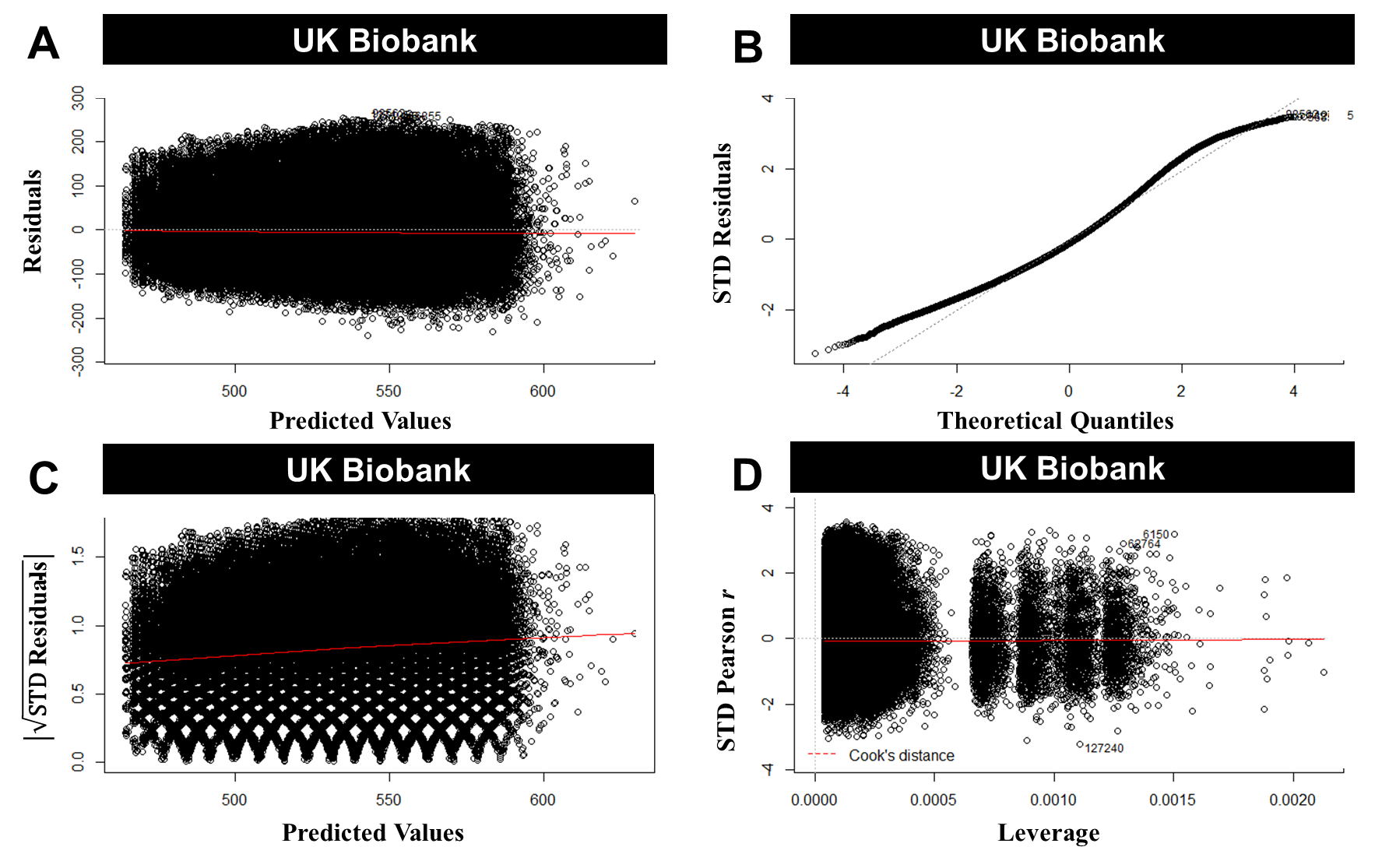
 **Supplementary Figure 5.** Regression diagnostic plots of the general linear model (LM) for the UK Biobank analyses (*n =* 157,933). (A) A plot of the residuals versus fitted values. This plot suggests that the assumptions of linearity, equality of variances, and no outliers are met. (B) Q-Q plot displaying the quantiles of the data versus quantiles of a normal distribution. This plot suggests there is an approximately normal distribution of the residuals and, thus, the error terms. (C) A plot of the absolute value of the standardized (STD) residuals’ square root versus the fitted values. This plot suggests that variability in residuals does not change much over the range of the dependent variable. (D) A plot of the residuals versus leverage. This plot shows that there are no influential cases, as all cases fall within Cook’s distance.

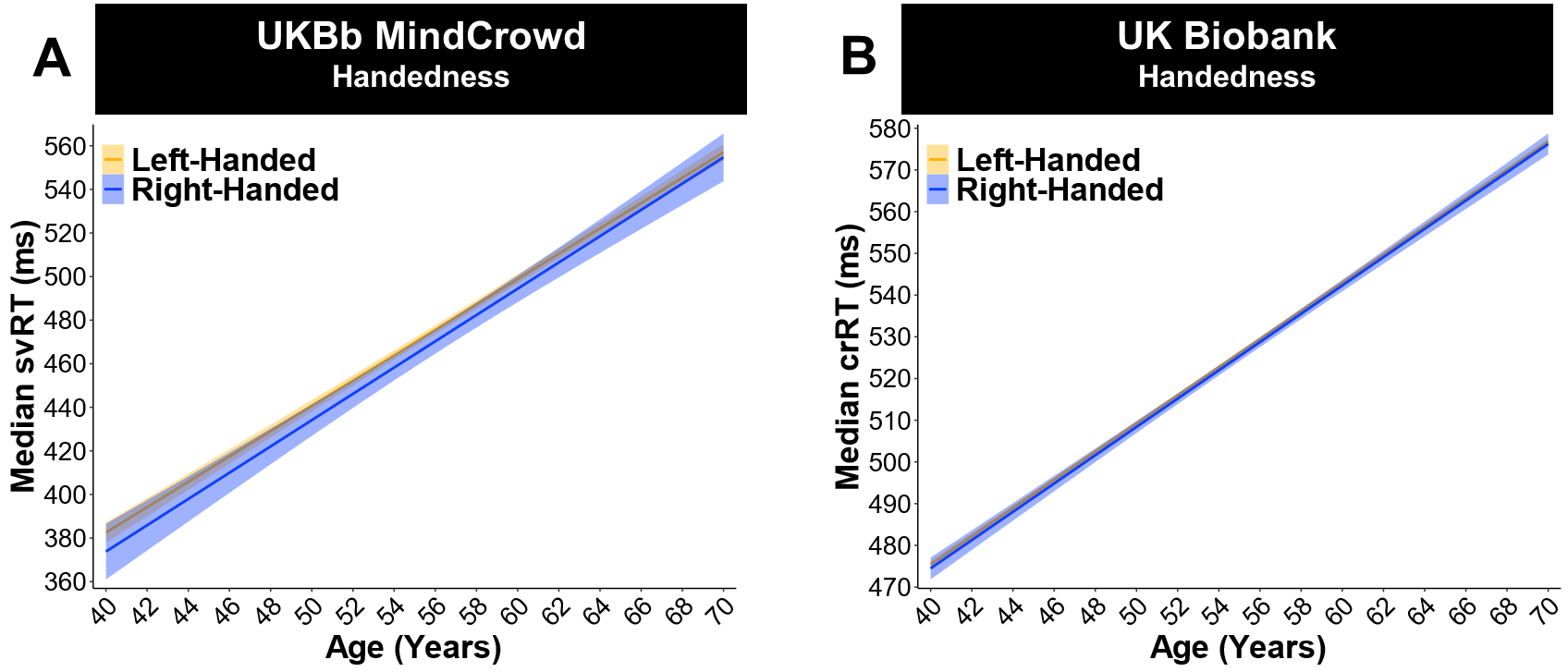

**Supplementary Figure 6.** Handedness was not associated with reaction time (RT) in the UK Biobank analysis (ages 40-70). (A) Linear model fits (line fill ± 95% CI) of median simple visual RT (svRT). There was no relationship between reported handedness and svRT performance in the UKBb MindCrowd cohort. (B) Linear model fits (line fill ± 95% CI) of median complex, visual, recognition RT (cvrRT). For both the UKBb MindCrowd *β_Handedness_* = -2.58, *p_Handedness_* = 4.00E-01, Left-Handed *n =* 4,520, Right-Handed *n =* 35,034), and the UK Biobank (*β_Handedness_* = 0.58, *p_Handedness_* = 0.36, Left-Handed *n =* 15,287, Right-Handed *n =* 142,958), handedness was not related to cvrRT in the UK Biobank.

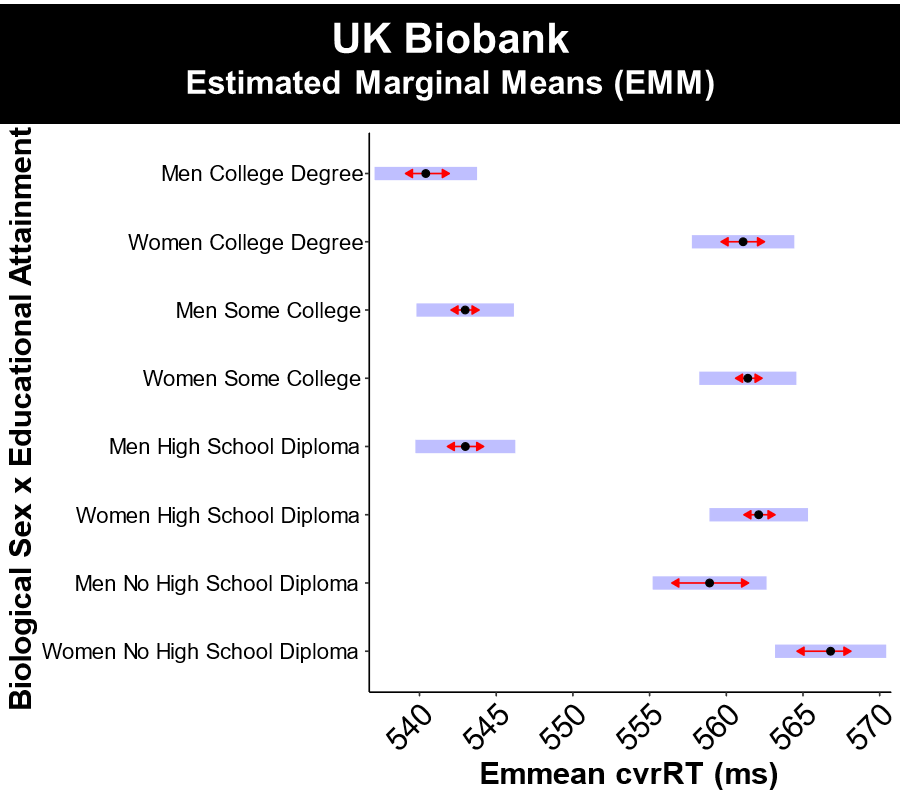
**Supplementary Figure 7.** Men with the lowest educational attainment show dramatically slower complex, visual, recognition reaction time (cvrRT). UK Biobank analysis (ages 40-70). Median cvrRT estimated marginal means (*EMM*) in the UK Biobank for the Biological Sex x Educational Attainment interaction. Purple/Blue rectangles display the range of the 95% confidence interval around the EMM, and the red arrows denote probable statistical significance if the arrows do not cross. Simple effects analysis via *EMM* revealed that men who did not graduate high school (*EMM* = 558.91 milliseconds), compared to men with more education (*EMMs* = 542.99,542.98, and 540.41 milliseconds), had markedly faster cvrRTs, more in line with the women’s cvrRT performance (*EMMs =* 566.80, 562.11, 561.40, and 561.09 milliseconds). In addition, the difference in cvrRT between men at the two lowest education milestones (*EMM* = 558.91 vs. 542.99 milliseconds*, β_Men_ =* 15.92, *p_Men_* = 2.00E-16, *n =* 4,922 and 18,505, 2.89%), was greater than those same two lowest milestones in women (*EMM* = 566.80 vs. 562.11 milliseconds*, β_Women_ =* 4.68, *p_Women_* = 3.12E-04, *n =* 6,056 and 27,742 , 0.84%). See Figure 14 for the Educational Attainment line graphs split by Biological Sex.

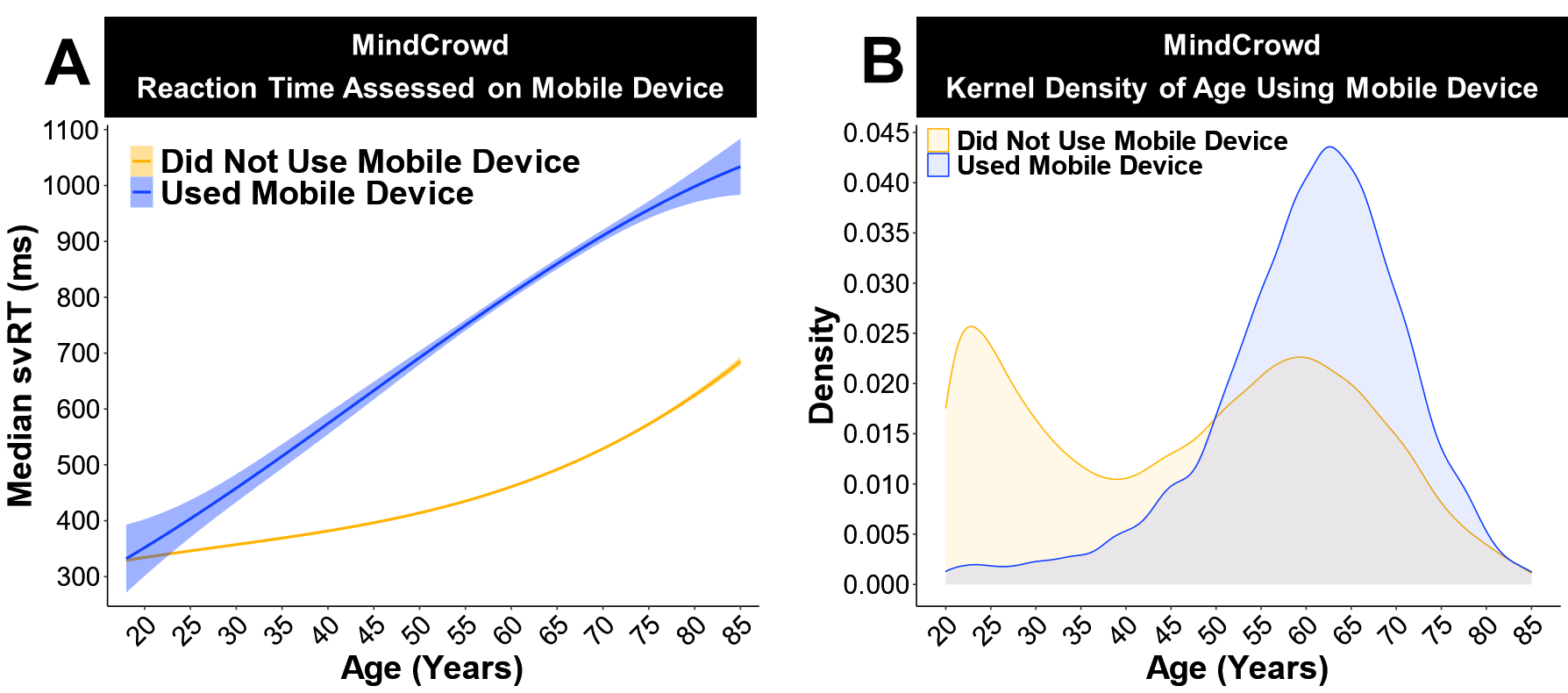

**Supplementary Figure 8.** Slower simple visual reaction time (svRT) when using a mobile device. MindCrowd analysis (ages 18-85): (A) linear model fits (line fill ± 95% *CI*) of the median svRT by Age^3^ (curvilinear model). Touch screen devices were related to markedly slower svRT performance across age (β*_Device_* = 322.30, *p_Device_* ≤ 2e-16, Used Mobile Device *n =* 7,603 age *M* = 54.06 *SD* = 14.54, Did Not Use Mobile Device *n =* 76,775 age *M* = 45.54 *SD* = 18.43). (B) Kernel density plot of the age distribution of participants using a mobile device. Participants using a mobile device. Those using a mobile device (i.e., using a touchscreen, *n* = 7,603, age *M* = 54.06 *SD* = 14.54 years) had slower svRTs and were older (β_Age~Mobile_ = 14.13, *p*_Age~Mobile_ < 2e-16) than those who did not use a mobile device. The MindCrowd svRT task procedure was as follows. Participants were presented with a pink sphere that appeared at random intervals (between 1-10s) on the screen, and they were instructed to respond as quickly as possible after the sphere appeared by pressing the enter or return key on their keyboard. Once the participant responded, the sphere disappeared until the next trial. Each participant received a total of five trials. The sphere stayed on the screen until the participant responded. The dependent variable, the response time (milliseconds), was recorded from the sphere’s appearance on the screen to the participant’s key press or screen touch.

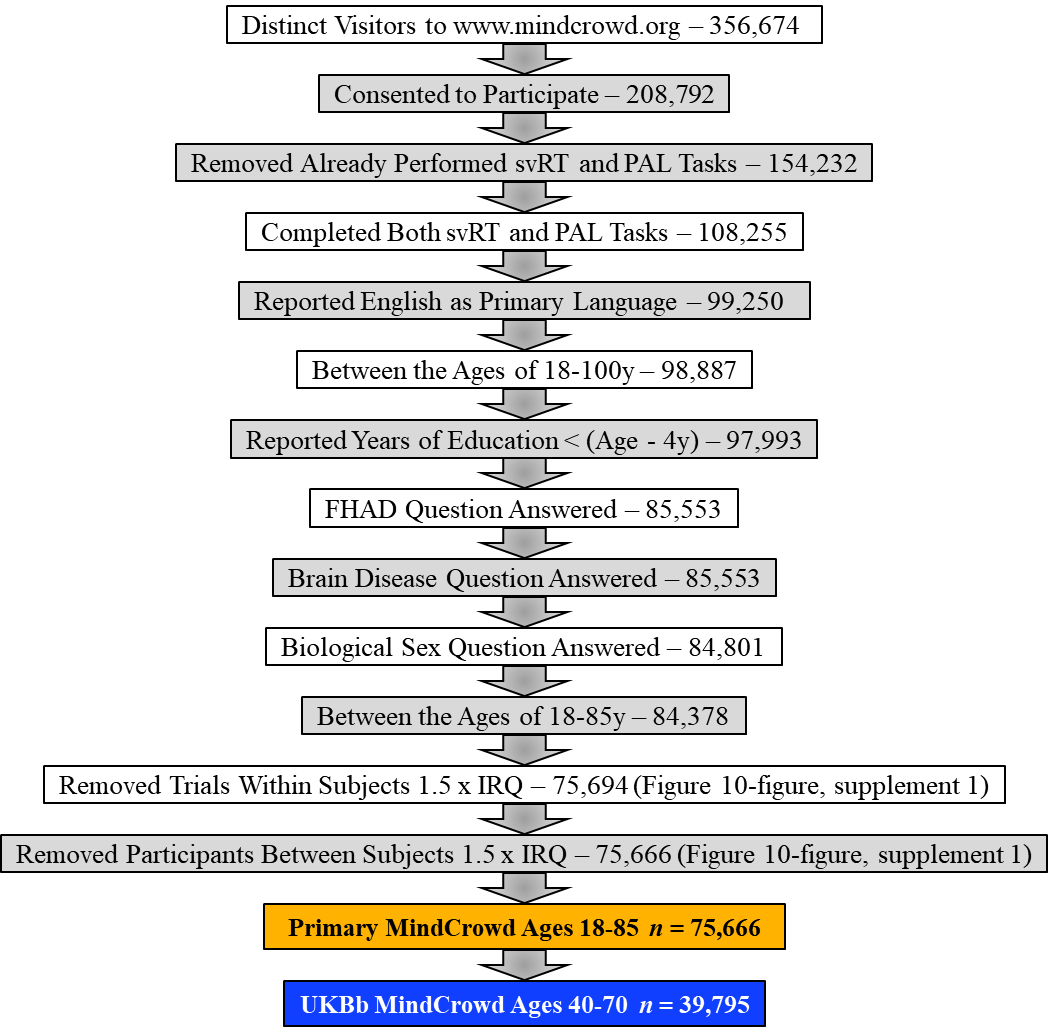

**Supplementary Figure 9.** Flowchart describing the data quality and control filtering steps, with associated change in *n* for each successive filtering step. This chart illustrates the primary filtering procedures used to ensure data quality for MindCrowd’s online evaluation and self-report.

**Supplementary Figure 10.** Identification of simple visual reaction time (svRT) outliers and the effects of their removal. (A) The distribution of median svRT by Age in the entire MindCrowd cohort (*n =* 84,378). This graph displays outliers (svRT ± 1.5 x interquartile range [*IQR*] and age > 85y) shaded in gray. (B) A plot of the distribution of median svRT by Age once svRT outliers were removed (*n =* 75,666). (C) A plot of the standard deviation (*SD*) of median svRT by Age in the entire MindCrowd cohort with outliers (svRT ± 1.5 x *IQR*) present. (D) Graph of the *SD* of median svRT by Age in the MindCrowd cohort once outliers were removed. This graph illustrates the direct effect of outliers staying above or below 1.5 x IQR of these data’s variation.
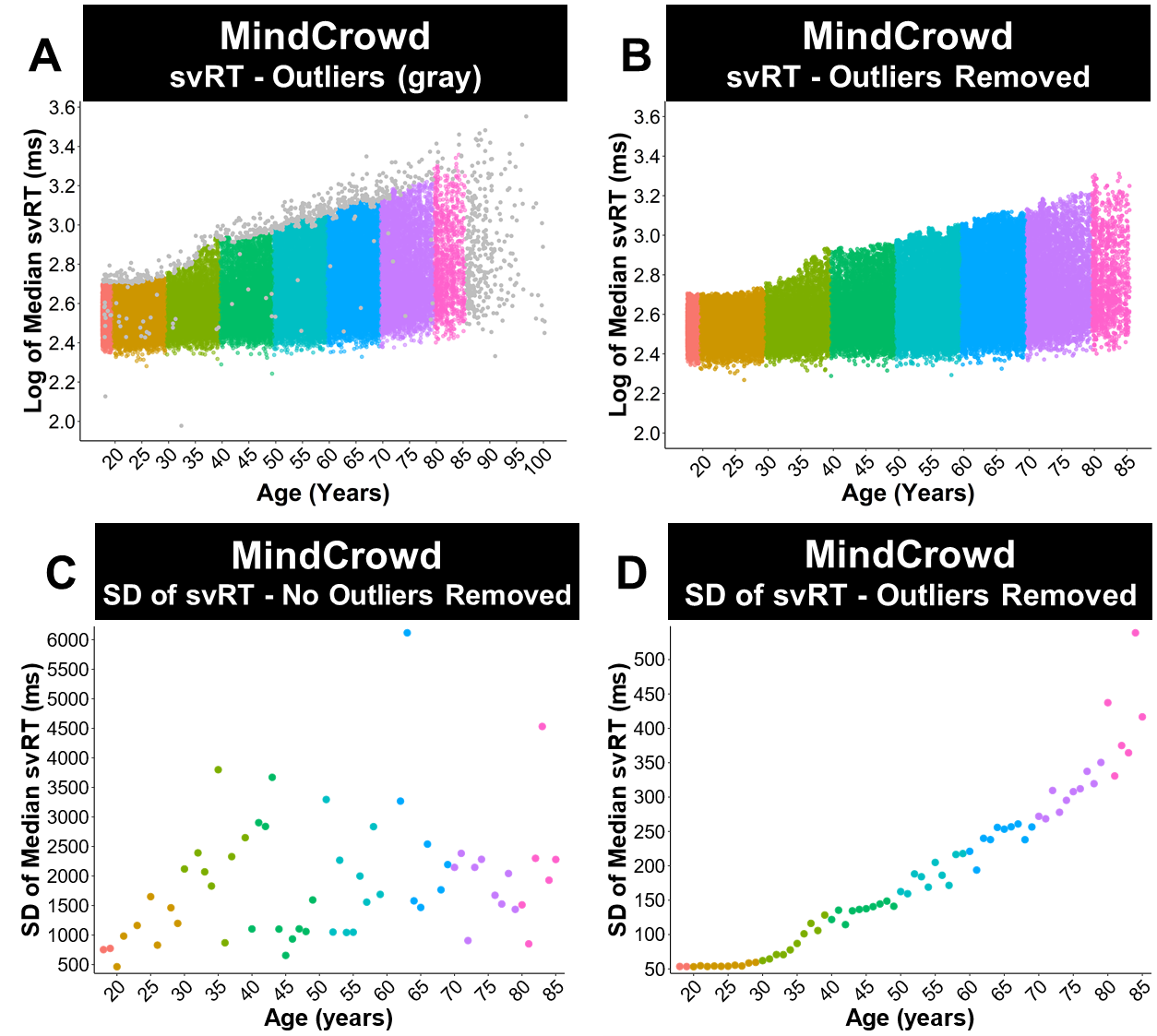

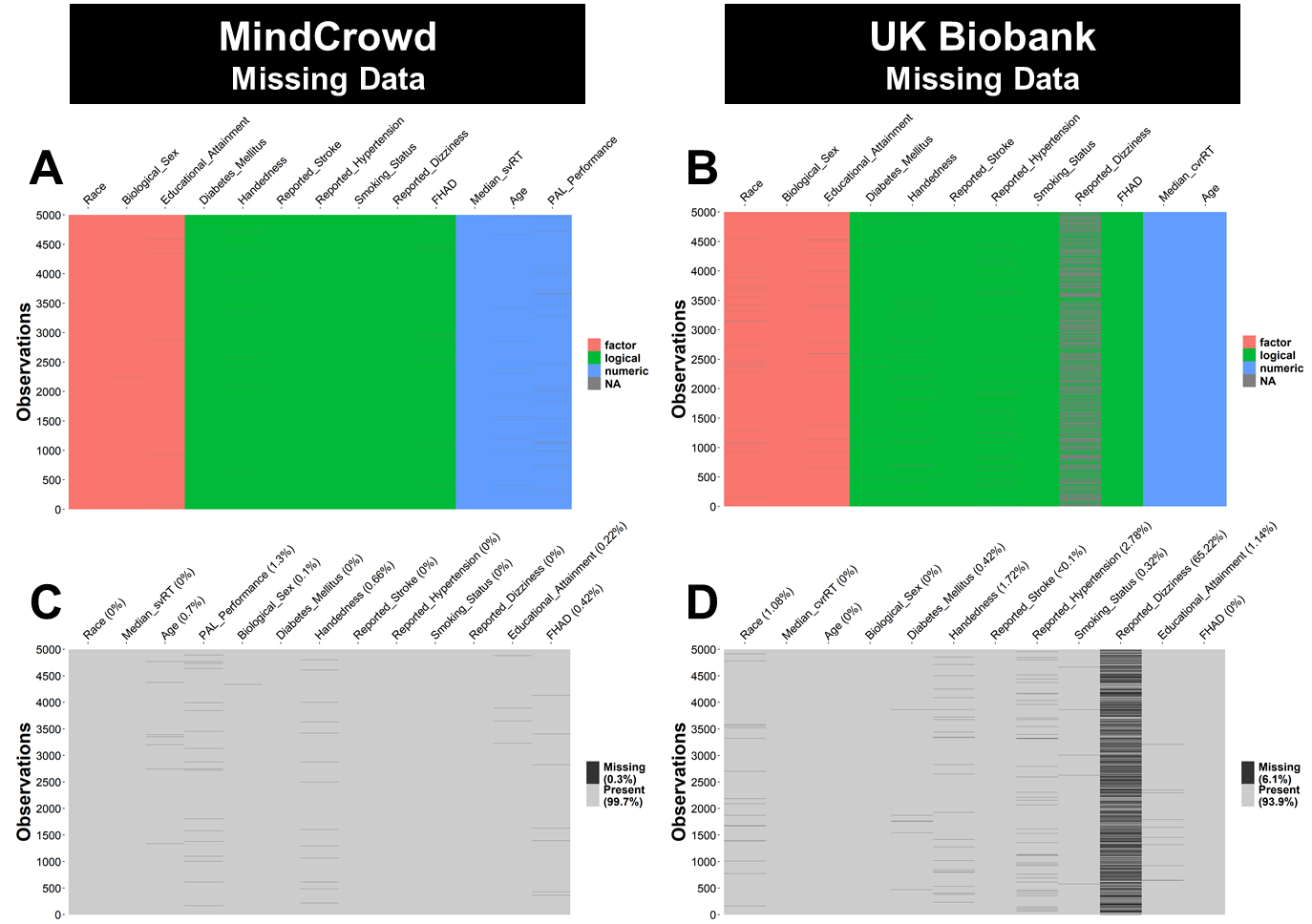
**Supplementary Figure 11.** Missing Data Patterns and Percentages MindCrowd and the UK Biobank. The plot area of each graph displays 5000 random selected cases from a total *n* = 97,967 for (A-C) MindCrowd (ages 18-85) and an *n* = 201,139 for (B-D) the UK Biobank (ages 40-70). (A-B) Columns colored according to a factor’s “Class.” Gray (NA) denotes dropped or missing data. All cases with NAs were listwise deleted before analysis. (C-D) Display the overall study’s “percent missing” and across each key factor. These percentages are derived from all the dataset cases, not the 5000 randomly selected to curtail graph size. The percent missing across both study cohorts and each key factor aside from Reported Dizziness in the UK Biobank was less than 5% (*M* = 0.50% *SD* = 0.74%). Reported Dizziness in the UK Biobank had 64.36% missing data. Thus, the scope of the interpretation of this factor’s significant association with reaction time (RT) is restricted (see Opportunities and Limitations of Our Research in the Discussion). The mean percent missing for the one unique and 12 shared key factors in MindCrowd was less than 1% (*M* = 0.31%). ). The mean percent missing for the 12 key shared factors in the UK Biobank was 7% (*M* = 7.04%). This average percent missing drops to less than 1% (*M* = 0.68%) when Reported Dizziness is excluded from the analysis.

| Supplementary Table 1 | | | |
| --- | --- | --- | --- |
| *Key Resources* | | | |
| Reagent Type,  Species, or Resource | **Designation** | **Source or Reference** | **Identifiers** |
| Software, algorithm | R | The R Foundation | Version 4.0.3 RRID: SCR_001905 |
| Software, algorithm | R package, tidyverse | Comprehensive R Archive Network (CRAN) | Version 1.3.0 RRID: SCR_014601: for ggplot2 |
| Software, algorithm | R package, glmulti | Comprehensive R Archive Network (CRAN) | Version 1.0.7.1 |
| Software, algorithm | R package, interactions | Comprehensive R Archive Network (CRAN) | Version 1.1.3 |
| Software, algorithm | R package, sandwich | Comprehensive R Archive Network (CRAN) | Version 2.5-1 |
| Software, algorithm | R package, jtools | Comprehensive R Archive Network (CRAN) | Version 2.0.5 |
| Software, algorithm | R package, emmeans | Comprehensive R Archive Network (CRAN) | Version 1.4.6 |
| Software, algorithm | RStudio | RStudio, PBC | Version 1.3.1093  RRID:SCR_000432 |
| Software, algorithm | R package, finalfit | Comprehensive R Archive Network (CRAN) | Version 1.0.2 |
| Software, algorithm | R package, visdat | Comprehensive R Archive Network (CRAN) | Version 0.5.3 |
| Software, algorithm | R package, naniar | Comprehensive R Archive Network (CRAN) | Version 0.6.0 |
| Software, algorithm | R package, qPCR | Comprehensive R Archive Network (CRAN) | Version 1.4-1 |

**Supplementary Table 1.** Key Resources. Table displaying critical reagents, software, and other items used in this study.

| Supplementary Table 2 | | |
| --- | --- | --- |
| ***MindCrowd: Participant Questions and Its Code Used for Analysis*** | | |
| **Question Number** | **Question Asked** | **Coded for Analysis** |
| Pre-Test, Question 1 | Sex: | Male, Female |
| Pre-Test, Question 2 | Age: | Numeric Free Response |
| Pre-Test, Question 3 | Primary Language: | 1-54 (the number assigned to one of 54 major languages) |
| Pre-Test, Question 4 | Education: | No High School Diploma |
|  |  | High School Diploma |
|  |  | Some College |
|  |  | College Degree |
| Pre-Test, Question 5 | Country: | ISO 3166 international country code (e.g., TZ, UA, UG, US) |
| Post-Test, Question 1 | What is your marital status? | Single, Married, Widowed, Unreported |
| Post-Test, Question 2 | Are you left or right-handed? | True, False |
| Post-Test, Question 3 | What is your Race? | American Indian or Alaska Native, Asian, Black/African American, Native Hawaiian/Pacific Islander, White, Mixed |
| Post-Test, Question 4 | Are you of Hispanic, Latino, or Spanish origin? | True, False |
| Post-Test, Question 5 | How many prescription medications do you take on a daily basis? | 0, 1, 2, 3, 4 |
| Post-Test, Question 6 | Please check all the following that apply to you. |  |
| 6a | Loss of consciousness ( > 10 minutes) | True, False |
| 6b | Seizures | True, False |
| 6c | Dizzy Spells | True, False |
| 6d | High Blood Pressure | True, False |
| 6e | Smoking Status | True, False |
| 6f | Diabetes | True, False |
| 6g | Heart Disease | True, False |
| 6h | Cancer | True, False |
| 6i | Reported Stroke | True, False |
| 6j | Alcohol/Drug Abuse | True, False |
| 6k | Brain Disease and/or Memory Problems | True, False |
| Post-Test, Question 7 | Have you, a sibling, or one of your parents been diagnosed with Alzheimer’s disease? | True, False, Not Recorded |

**Supplementary Table 2.** Health, Medical, Lifestyle, and Demographic Questions. Table displaying questions participants were asked on the MindCrowd website. Other information for each question includes if it was asked per or post-testing, its number, and how it was coded for statistical analysis.

| Supplementary Table 3 | | | | | | | |
| --- | --- | --- | --- | --- | --- | --- | --- |
| ***MindCrowd, UKBb MindCrowd, and the UK Biobank Variance Inflation Factors (VIF)*** | | | | | | | |
| **Factor** | ***df*** | **MindCrowd** | | **UKBb MindCrowd** | | **UK Biobank** | |
|  |  | VIF | VIF^(1/(2*Df))^ | VIF | VIF^(1/(2*^*^df^*^))^ | VIF | VIF^(1/(2*^*^df^*^))^ |
| Age^1^ | 1 | 1076.88 | 32.82 | 1.49 | 1.22 | 1.95 | 1.40 |
| Age^2^ | 1 | 3491.06 | 59.09 |  |  |  |  |
| Age^3^ | 1 | 961.20 | 31.00 |  |  |  |  |
| PAL Performance^1^ | 1 | 159.95 | 12.65 |  |  |  |  |
| PAL Performance^2^ | 1 | 751.04 | 27.41 |  |  |  |  |
| PAL Performance^3^ | 1 | 275.83 | 16.61 |  |  |  |  |
| Biological Sex | 1 | 7.81 | 2.79 | 106.20 | 10.31 | 78.48 | 8.86 |
| Educational Attainment | 3 | 231.58 | 2.48 | 2.45 | 1.16 | 6.06 | 1.35 |
| Handedness | 1 | 1.00 | 1.00 | 1.00 | 1.00 | 1.00 | 1.00 |
| Daily Medications Taken | 4 | 1.96 | 1.09 |  |  |  |  |
| Reported Dizziness | 1 | 1.06 | 1.03 | 1.01 | 1.01 | 1.01 | 1.01 |
| Smoking Status | 1 | 7.60 | 2.76 | 1.03 | 1.02 | 1.03 | 1.01 |
| Diabetes Mellitus | 1 | 1.16 | 1.08 | 1.06 | 1.03 | 1.03 | 1.02 |
| Reported Stroke | 1 | 23.94 | 4.89 | 1.01 | 1.01 | 1.01 | 1.00 |
| Reported Hypertension | 1 | 1.37 | 1.17 | 1.10 | 1.05 | 1.08 | 1.04 |
| FHAD | 1 | 1.13 | 1.06 | 1.02 | 1.01 | 1.02 | 1.01 |
| Age x PAL Performance | 1 | 11.97 | 3.46 |  |  |  |  |
| Age x Smoking Status | 1 | 7.40 | 2.72 |  |  |  |  |
| Age x Reported Stroke | 1 | 23.95 | 4.89 |  |  |  |  |
| Age x Biological Sex | 1 | 7.46 | 2.73 | 45.48 | 6.74 | 57.39 | 7.58 |
| Age x Educational Attainment | 3 | 2100.05 | 3.58 | 104.10 | 2.17 | 50.24 | 1.92 |

**Supplementary Table 3.** Summary of the variance inflation factors (VIF) across the three cohorts. These data suggest that the VIF for most factors was below conventional “high” multicollinearity levels (i.e., 5). The factors with VIF > 5 were expected to show higher values (e.g., polynomials, interaction terms, and dummy variables > three levels) and thus should not have meaningfully impacted model results.

| Supplementary Table 4 | | | |
| --- | --- | --- | --- |
| ***MindCrowd Missing Data Information*** | | | |
| **Factor** | **Factor *n*** | **Missing *n*** | **Percent Missing** |
| Age | 96313 | 674 | 0.69 |
| PAL Performance | 95583 | 1404 | 1.45 |
| Biological Sex | 96916 | 71 | 0.07 |
| Race | 96987 | 0 | 0.00 |
| Diabetes Mellitus | 96987 | 0 | 0.00 |
| Handedness | 96436 | 551 | 0.57 |
| Reported Stroke | 96987 | 0 | 0.00 |
| Reported Hypertension | 96987 | 0 | 0.00 |
| Smoking Status | 96987 | 0 | 0.00 |
| Reported Dizziness | 96987 | 0 | 0.00 |
| Educational Attainment | 96691 | 296 | 0.31 |
| FHAD | 96589 | 398 | 0.41 |

**Supplementary Table 4.** Missing data numbers for MindCrowd. The Ten key factors in MindCrowd shared by the UK Biobank had relatively little missing data (merged reported and excluded *n =* 96,987, Missing Data Percentage *M* = 0.32% *SD* = 0.45%). Table acronym: A first-degree family history of Alzheimer’s disease (FHAD).

| Supplementary Table 5 | | | |
| --- | --- | --- | --- |
| ***UK Biobank Missing Data Information*** | | | |
| **Factor** | **Factor *n*** | **Missing *n*** | **Percent Missing** |
| Age | 201139 | 0 | 0 |
| Biological Sex | 201139 | 0 | 0 |
| Race | 198737 | 2402 | 1.2 |
| Diabetes Mellitus | 200511 | 628 | 0.31 |
| Handedness | 197525 | 3614 | 1.79 |
| Reported Stroke | 201040 | 99 | < 0.00 |
| Reported Hypertension | 195867 | 5272 | 2.62 |
| Smoking Status | 200411 | 728 | 0.36 |
| Reported Dizziness | 71676 | 129463 | 64.36 |
| Educational Attainment | 199127 | 2012 | 1.00 |
| FHAD | 201139 | 0 | 0 |

**Supplementary Table 5.** Missing data numbers for the UK Biobank. Nine out of the ten key factors in the UK Biobank shared by MindCrowd had relatively little missing data (merged reported and excluded *n =* 201,139, *M* = 7.56%. *SD* = 17.65%). One potentially concerning factor in the UK Biobank was Reported Dizziness with 64.36% missing. Reported Dizziness had a significant association with cvrRT in our linear model. Interpretation of Reported Dizziness’ association with complex visual recognition reaction time (cvrRT) should only be used for “hypothesis generation.” Table acronym: A first-degree family history of Alzheimer’s disease (FHAD).

| Supplementary Table 6 | | | | | | | | | |
| --- | --- | --- | --- | --- | --- | --- | --- | --- | --- |
| ***MindCrowd: Odds Ratios (95% CIs) for Exclusion Due to Missing or Removed Data*** | | | | | | | | | |
| **Factor** | **Factor Level** | **Reported** | | | **Excluded** | | | **OR** | **95% CI** |
|  |  | ***n*** | **%** | **Odds** | ***n*** | **%** | **Odds** |  |  |
| Age Decade | 18-20 | 289 | 10.38 | 0.62 | 142 | 3.99 | 0.24 | 0.38 | (0.48, 0.30) |
|  | 20-30 | 796 | 28.58 | 1.71 | 262 | 7.36 | 0.44 | 0.26 | (0.31, 0.21) |
|  | 30-40 | 398 | 14.29 | 0.86 | 192 | 5.39 | 0.32 | 0.38 | (0.46, 0.30) |
|  | 40-50 | 384 | 13.79 | 0.83 | 291 | 8.17 | 0.49 | 0.59 | (0.72, 0.49) |
|  | ***50-60^a^*** | ***465*** | ***16.70*** |  | ***597*** | ***16.77*** |  |  |  |
|  | 60-70 | 343 | 12.32 | 0.74 | 587 | 16.49 | 0.98 | 1.33 | (1.60, 1.11) |
|  | 70-80 | 100 | 3.59 | 0.22 | 323 | 9.07 | 0.54 | 2.52 | (3.25, 1.95) |
|  | 80-90 | 10 | 0.36 | 0.02 | 72 | 2.02 | 0.12 | 5.61 | (10.99, 2.86) |
| Biological Sex | Women | 1,817 | 65.24 | 1.88 | 2272 | 63.82 | 1.87 | 0.99 | (1.10, 0.90) |
|  | Men | 968 | 34.76 |  | 1217 | 34.19 |  |  |  |
| Race | Asian | 1,570 | 56.37 | 3.51 | 229 | 6.43 | 0.08 | 0.02 | (0.03, 0.02) |
|  | Black | 497 | 17.85 | 1.11 | 165 | 4.63 | 0.06 | 0.05 | (0.06, 0.04) |
|  | Mixed | 271 | 9.73 | 0.61 | 300 | 8.43 | 0.10 | 0.17 | (0.21, 0.14) |
|  | ***White^a^*** | ***447*** | ***16.05*** |  | ***2866*** | ***80.51*** |  |  |  |
| Educational Attainment | ***No High School Diploma^a^*** | ***137*** | ***4.92*** |  | ***507*** | ***14.24*** |  |  |  |
|  | High School Diploma | 340 | 12.21 | 2.48 | 438 | 12.30 | 0.86 | 0.35 | (0.44, 0.27) |
|  | Some College | 1,070 | 38.42 | 7.81 | 798 | 22.42 | 1.57 | 0.20 | (0.25, 0.16) |
|  | College Degree | 1,238 | 44.45 | 9.04 | 1521 | 42.72 | 3.00 | 0.33 | (0.41, 0.27) |
| Diabetes Mellitus | FALSE | 2,574 | 92.42 | 0.08 | 3308 | 92.92 | 0.08 | 0.93 | (1.12, 0.77) |
|  | TRUE | 211 | 7.58 |  | 252 | 7.08 |  |  |  |
| Handedness | FALSE | 2,412 | 86.61 | 0.15 | 2670 | 75.00 | 0.13 | 0.87 | (1.01, 0.74) |
|  | TRUE | 354 | 12.71 |  | 339 | 9.52 |  |  |  |
| Reported Stroke | FALSE | 2,734 | 98.17 | 0.02 | 3495 | 98.17 | 0.02 | 1.00 | (0.69, 0.69) |
|  | TRUE | 51 | 1.83 |  | 65 | 1.83 |  |  |  |
| Reported Hypertension | FALSE | 2,189 | 78.60 | 0.27 | 2768 | 77.75 | 0.29 | 1.05 | (1.19, 0.93) |
|  | TRUE | 596 | 21.40 |  | 792 | 22.25 |  |  |  |
| Smoking Status | FALSE | 2,466 | 88.55 | 0.13 | 3235 | 90.87 | 0.10 | 0.78 | (0.91, 0.66) |
|  | TRUE | 319 | 11.45 |  | 325 | 9.13 |  |  |  |
| Reported Dizziness | FALSE | 2,534 | 90.99 | 0.10 | 3178 | 89.27 | 0.12 | 1.21 | (1.43, 1.03) |
|  | TRUE | 251 | 9.01 |  | 382 | 10.73 |  |  |  |
| FHAD | FALSE | 2,198 | 78.92 | 0.27 | 2385 | 66.99 | 0.33 | 1.22 | (1.38, 1.08) |
|  | TRUE | 587 | 21.08 |  | 777 | 21.83 |  |  |  |

**Supplementary Table 6.** Odds ratios (OR) with 95% CI for evaluation of selection bias within MindCrowd. Being excluded due to missing data (i.e., via listwise deletion) had a lower likelihood across most key factor levels. Exceptions were participants over 60 years old that had a higher likelihood of being excluded. In addition, participants reporting hypertension, dizziness, and a first-degree family history of Alzheimer’s disease (FHAD) also had a higher probability of being excluded. ***^a^*** denotes the factor level used as a reference for odds calculations.

| Supplementary Table 7 | | | | | | | | | |
| --- | --- | --- | --- | --- | --- | --- | --- | --- | --- |
| ***The UK Biobank: Odds Ratios (95% CIs) for Exclusion to Missing or Removed Data*** | | | | | | | | | |
| **Factor** | **Factor Level** | **Reported** | | | **Excluded** | | | **OR** | **95% CI** |
|  |  | ***n*** | **%** | **Odds** | ***n*** | **%** | **Odds** |  |  |
| Age Decade | 40-50 | 38,167 | 24.12 | 0.64 | 30961 | 23.27 | 0.75 | 1.18 | (1.20, 1.16) |
|  | ***50-60^a^*** | ***59,904*** | ***37.85*** |  | ***41202*** | ***30.97*** |  |  |  |
|  | 60-70 | 59,721 | 37.74 | 1.00 | 59952 | 45.06 | 1.46 | 1.46 | (1.48, 1.44) |
| Biological Sex | Women | 89,333 | 56.45 | 1.30 | 62484 | 46.96 | 0.89 | 0.68 | (0.69, 0.67) |
|  | Men | 68,918 | 43.55 |  | 70571 | 53.04 |  |  |  |
| Race | Asian | 847 | 0.54 | 0.01 | 777 | 0.58 | 0.01 | 1.31 | (1.47, 1.19) |
|  | Black | 154,812 | 97.83 | 1.73 | 124003 | 93.20 | 1.98 | 1.15 | (1.22, 1.13) |
|  | Mixed | 980 | 0.62 | 0.01 | 2465 | 1.85 | 0.02 | 3.14 | (3.45, 2.91) |
|  | ***White^a^*** | ***1,612*** | ***1.02*** |  | ***3408*** | ***2.56*** |  |  |  |
| Educational Attainment | ***No High School Diploma^a^*** | ***10,979*** | ***6.94*** |  | ***28149*** | ***21.16*** |  |  |  |
|  | High School Diploma | 46,248 | 29.22 | 4.21 | 37312 | 28.04 | 1.33 | 0.31 | (0.32, 0.31) |
|  | Some College | 77,272 | 48.83 | 7.04 | 52680 | 39.59 | 1.87 | 0.27 | (0.27, 0.26) |
|  | College Degree | 23,752 | 15.01 | 2.16 | 12902 | 9.70 | 0.46 | 0.21 | (0.22, 0.21) |
| Diabetes Mellitus | FALSE | 153,281 | 96.86 | 0.03 | 124202 | 93.35 | 0.07 | 2.04 | (2.12, 2.01) |
|  | TRUE | 4,970 | 3.14 |  | 8225 | 6.18 |  |  |  |
| Handedness | FALSE | 142,964 | 90.34 | 0.11 | 117170 | 88.06 | 0.10 | 0.98 | (1.00, 0.97) |
|  | TRUE | 15,287 | 9.66 |  | 12271 | 9.22 |  |  |  |
| Reported Stroke | FALSE | 157,013 | 99.22 | 0.01 | 130649 | 98.19 | 0.02 | 2.24 | (2.40, 2.21) |
|  | TRUE | 1,238 | 0.78 |  | 2307 | 1.73 |  |  |  |
| Reported Hypertension | FALSE | 125,655 | 79.40 | 0.26 | 93491 | 70.26 | 0.37 | 1.41 | (1.44, 1.39) |
|  | TRUE | 32,596 | 20.60 |  | 34292 | 25.77 |  |  |  |
| Smoking Status | FALSE | 63,953 | 40.41 | 1.47 | 52371 | 39.36 | 1.53 | 1.04 | (1.05, 1.02) |
|  | TRUE | 94,298 | 59.59 |  | 79956 | 60.09 |  |  |  |
| Reported Dizziness | FALSE | 116,040 | 73.33 | 0.36 | 2466 | 1.85 | 0.46 | 1.26 | (1.35, 1.20) |
|  | TRUE | 42,211 | 26.67 |  | 1126 | 0.85 |  |  |  |
| FHAD | FALSE | 138,509 | 87.52 | 0.14 | 118091 | 88.75 | 0.13 | 0.89 | (0.91, 0.88) |
|  | TRUE | 19,742 | 12.48 |  | 14964 | 11.25 |  |  |  |

**Supplementary Table 7.** Odds ratios (OR) with 95% CI for evaluation of selection bias within the UK Biobank. The odds of being excluded due to incomplete cases with missing data. The odds of being excluded due to missing data (i.e., listwise deletion) was more likely across over half of the key factor levels. Notable factor levels with a greater probability of being excluded were participants over 60 years old and those identified as Asian or Mixed race. In addition, participants reporting diabetes, stroke, hypertension, and dizziness also had a higher likelihood of being excluded. ***^a^*** denotes the factor level used as a reference for odds calculations.
